# Genome-wide meta-analysis and omics integration identifies novel genes associated with diabetic kidney disease

**DOI:** 10.1101/2021.08.27.21262264

**Authors:** Niina Sandholm, Joanne B Cole, Viji Nair, Xin Sheng, Hongbo Liu, Emma Ahlqvist, Natalie van Zuydam, Emma H Dahlström, Damian Fermin, Laura J Smyth, Rany M Salem, Carol Forsblom, Erkka Valo, Valma Harjutsalo, Eoin P Brennan, Gareth McKay, Darrell Andrews, Ross Doyle, Helen C Looker, Robert G Nelson, Colin Palmer, Amy Jayne McKnight, Catherine Godson, Alexander P Maxwell, Leif Groop, Mark I McCarthy, Matthias Kretzler, Katalin Susztak, Joel N Hirschhorn, Jose C Florez, Per-Henrik Groop, for the GENIE Consortium

**Author notes:** Authors contributed equally to this work (Niina Sandholm, Joanne B Cole).

## Abstract

**Background:** Diabetes is the leading cause of kidney disease, and heritability studies demonstrate a substantial, yet poorly understood, contribution of genetics to kidney complications in people with diabetes.

**Methods:** We performed genome-wide association study (GWAS) meta-analyses using ten different phenotypic definitions of diabetic kidney disease (DKD), including nearly 27,000 individuals with diabetes, and integrated the results with various kidney omics datasets.

**Results:** The meta-analysis identified a novel low frequency intronic variant (rs72831309) in the *TENM2* gene encoding teneurin transmembrane protein 2 associated with a lower risk of the combined chronic kidney disease (CKD; eGFR<60 ml/min/1.73 m^2^) and DKD (microalbuminuria or worse) phenotype (“CKD-DKD”, odds ratio 2.08, *p*=9.8×10^−9^). Gene-level analysis identified ten genes associated with DKD (*COL20A1, DCLK1, EIF4E, PTPRN-RESP18, GPR158, INIP-SNX30, LSM14A*, and *MFF, p*<2.7×10^−6^). Integration of GWAS data with human glomerular and tubular expression data in a transcriptome-wide association study demonstrated higher tubular *AKIRIN2* gene expression in DKD versus non-DKD controls (*p*=1.1×10^−6^). The lead SNPs within the *DCLK1, AKIRIN2, SNX30* and three other gene regions significantly alterated the methylation at this region in kidneys (*p*<2.2×10^−11^). Expression of target genes in kidney tubules or glomeruli correlated with relevant pathological phenotypes. For example, tubular *TENM2* expression positively correlated with eGFR (*p*=2.3×10^−9^) and negatively with tubulointerstitial fibrosis (*p*=4.7×10^−9^), tubular *DCLK1* expression positively correlated with fibrosis (*p*=1.6×10^−12^), and *SNX30* level positively correlated with eGFR (*p*=7.6×10^−13^) and negatively with fibrosis (*p*<2×10^−16^).

**Conclusions:** GWAS meta-analysis and integration with renal omics data points to novel genes contributing to pathogenesis of DKD.

## INTRODUCTION

Diabetes is the leading cause of kidney disease. Diabetic kidney disease (DKD) is associated with high cardiovascular risk^1^ and mortality^2^, and consequently, both diabetes and kidney disease are among the most important causes of death worldwide^3^. While environmental factors, and especially blood glucose control, have a major impact on the risk of developing DKD, genetic factors also contribute to disease^4,5^. Although more than 300 genetic loci have been associated with chronic kidney disease (CKD) in the general population, these loci show limited effect in DKD, especially in individuals with type 1 diabetes (T1D)^6^. Genome-wide association studies (GWAS) have previously identified a handful of genetic loci for DKD in individuals with T1D at the genome-wide significance level (*p*<5×10^−8^)^7–10^. Recently, a GWAS meta-analysis including up to 19,406 individuals with T1D from the Diabetic Nephropathy Collaborative Research Initiative (DNCRI) identified 16 loci associated with various DKD definitions. The strongest association was a common missense variant on the *COL4A3* gene which also showed evidence of association in individuals with type 2 diabetes (T2D).^6^ A GWAS meta-analysis from the The SUrrogate markers for Micro- and Macrovascular hard endpoints for Innovative diabetes Tools (SUMMIT) consortium, including 6,000 individuals with T2D from five different studies, identified three loci for DKD in T2D, including the *UMOD* and *PRKAG2* loci previously identified in the general population^11^. However, meta-analysis with SUMMIT T1D studies (N=5,156)^4^ did not yield any genome-wide significant findings^11^. In this study, we performed GWAS meta-analyses on ten different DKD case-control definitions, including nearly 27,000 individuals with T1D or T2D from the two large consortia (including DNCRI^6^, SUMMIT-T1D^4^ and SUMMIT-T2D^11^ studies) to increase the power to detect novel genetic risk factors, followed by integration with versatile biological data to improve our understanding of the underlying biological mechanisms and clinical correlations (**Figure 1**).

**Figure 1:**
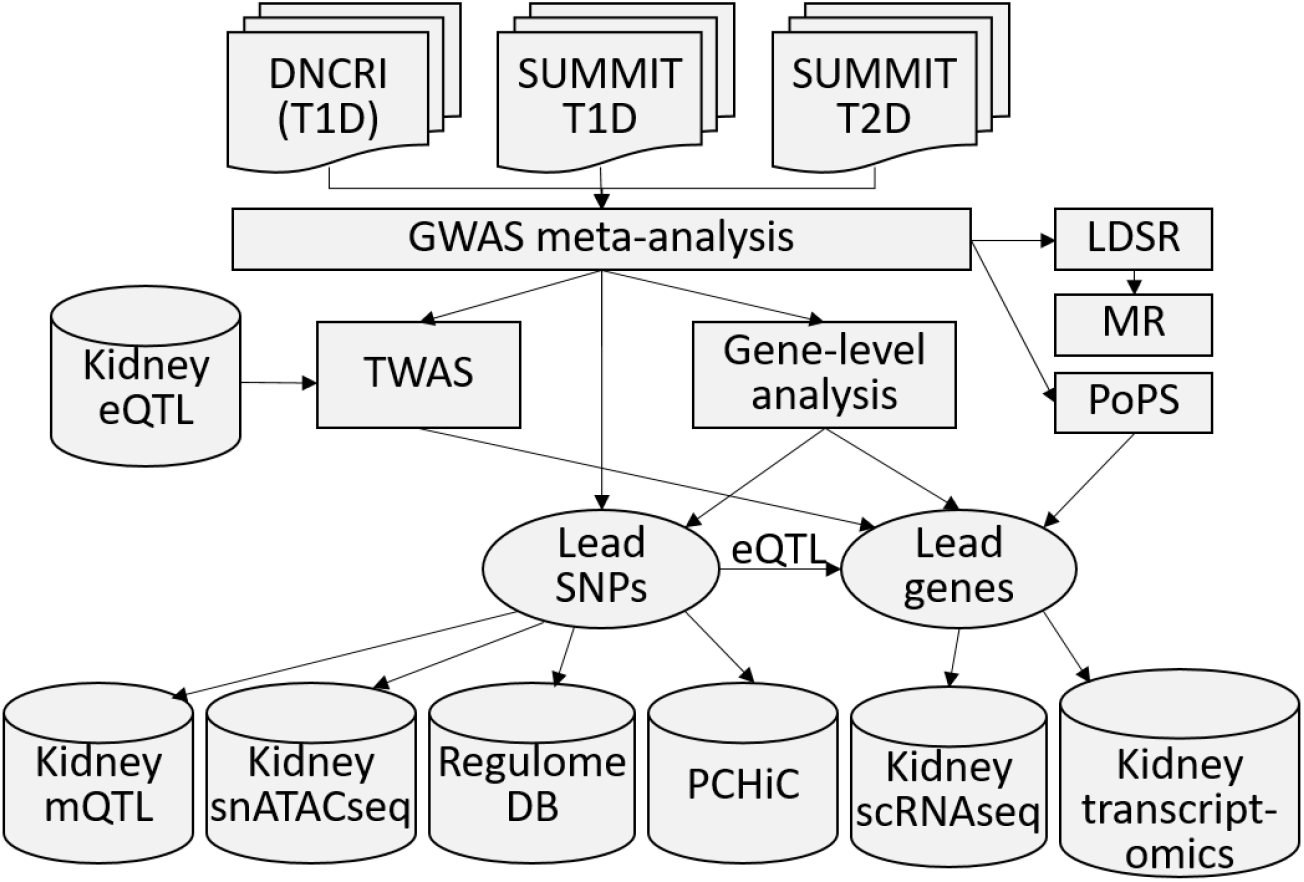
Schematic illustration of the study design from GWAS meta-analysis to integration with various omics data sets. GWAS meta-analysis for ten different phenotypic definitions of DKD included up to 26,785 individuals with either T1D or T2D from the previous DNCRI and SUMMIT GWAS meta-analyses. TWAS: Transcriptome-wide association study, integrating the GWAS meta-analysis results with kidney expression quantitative trait locus (eQTL) data for tubular and glomerular compartments, identifying genes with differential expression in DKD. mQTL: methylation quantitative trait locus (mQTL) data, identifying SNPs associated with DNA methylation at CpG sites. snATACseq: single nucleus ATAC sequencing, informative of chromatin openness in various kidney cell types. RegulomeDB: database with extensive epigenetic annotation for SNPs. PCHiC: promoter capture HiC sequencing data for sequence interaction with gene promoters, proposing target genes. Kidney transcriptomics: gene expression in glomerular and tubular tissue in nephrectomy samples, or in Pima Indian biopsies, correlated with various renal parameters.

## METHODS

### Participating studies and phenotype definitions

A total of ten case – control definitions for DKD were included in DNCRI^6^, based on either urinary albumin excretion rate (AER; divided into controls with normal AER, and cases with microalbuminuria, macroalbuminuria, or ESRD), estimated glomerular filtration rate (eGFR), or both, and harmonized to match and include all seven phenotypic definitions assessed in SUMMIT-T1D^4^ and SUMMIT-T2D^11^ analyses (**Supplemental Table 1**). All individuals (both cases and controls) had diabetes (either T1D or T2D) and cases had some form of kidney disease. The phenotypic comparisons are as follows: controls with normal AER vs. DKD cases with microalbuminuria or worse (“All vs. Ctrl”), macroalbuminuria or worse (“Severe DKD”), microalbuminuria alone (“Micro”), or “ESRD”; ESRD cases vs. everyone else (“ESRD vs. All”); controls with normal eGFR defined as eGFR ≥ 60 ml/min/1.73 m^2^ vs. CKD defined as eGFR < 60 ml/min/1.73 m^2^ (“CKD”); and “CKD-DKD” based on both AER and eGFR, with controls with normal AER and eGFR vs. cases with microalbuminuria or worse and eGFR < 45 ml/min/1.73 m^2^. For the three phenotypic comparisons not initially part of the SUMMIT analysis (normal AER vs. macroalbuminuria [“Macro”], ESRD vs. macroalbuminuria [“ESRD vs. macro”], and controls with eGFR ≥ 60 ml/min/1.73m^2^ vs. CKD cases with eGFR < 15 ml/min/1.73m^2^ or ESRD [“CKD extremes”]), GWAS and meta-analysis were performed with three SUMMIT T2D studies (Genetics of Diabetes Audit and Research, Tayside and Scotland [GoDARTS] 1 and 2, and Scania Diabetes Registry [SDR] T2D cohort) and the SDR T1D cohort. Individuals from the Finnish Diabetic Nephropathy Study (FinnDiane) were included in both the original DNCRI (N=6,019) and SUMMIT-T1D analyses (N=3,415), but for the purpose of this meta-analysis, FinnDiane was only included in the DNCRI meta-analysis and therefore excluded from the SUMMIT-T1D data (**Supplemental Table 2**). All contributing studies were performed in accordance with the Declaration of Helsinki and Declaration of Istanbul.

### Statistical analysis

Genotyping and statistical analysis of the DNCRI^6^ and SUMMIT^4,11^ cohorts have been previously described. Analysis plans were similar in both consortia, and the main characteristics are described in **Supplemental Table 3**. Imputation was performed using 1000Genomes Phase 3 reference panel in DNCRI, and the older 1000Genomes phase 1 panel in the SUMMIT cohorts. In both consortia, analyses were performed in unrelated individuals using the SNPtest additive score test, adjusting for age, sex, diabetes duration, the genetic principal components, and study-specific covariates (e.g., site or genotyping batch). Variants were filtered for INFO imputation quality score ≥ 0.3 (DNCRI) or ≥ 0.4 (SUMMIT) and minor allele count ≥ 10 in both cases and in controls. In SUMMIT, variants were further filtered to those with minor allele frequency (MAF) ≥ 0.01. Within-consortium meta-analyses were performed with inverse-variance fixed effects meta-analysis based on the effect size estimates. Meta-analyses between DNCRI, SUMMIT-T1D, and SUMMIT-T2D were performed with inverse-variance fixed effect methods based on the effect size estimates from the summary statistics for each of the three datasets. Finally, variants were limited to those found in at least two studies and reported in the 1000Genomes phase 3 reference panel. Study-wise summary statistics were available for DNCRI and SUMMIT-T1D studies. Regional association plots were plotted with LocusZoom.^12^

### Conditional analysis

We performed conditional analysis of the *COL4A3* locus with apparent secondary association peak using GCTA v1.93β ^13^ and FinnDiane GWAS data as the reference panel.

### Gene-level analysis

SNP summary statistics from the GWAS meta-analysis were aggregated by gene-level regression analysis using two related software programs, MAGMA^14^ and PASCAL^15^, using default parameters. Gene-level significance thresholds were determined by a Bonferroni multiple-testing correction based on the number of genes tested for each of the ten phenotypes within each software program (number of genes ranged from 18,439-21,790; significance thresholds ranged from 2.7×10^−6^ to 2.3×10^−6^).

### Transcriptome-wide association study (TWAS)

MetaXcan^16^ was applied to integrate kidney eQTL datasets with the GWAS meta-analysis results and to map disease-associated genes. The *cis-* eQTL data for microdissected human glomerular (N=119) and tubular (N=121) samples were obtained from Susztaklab Kidney Biobank (https://susztaklab.com/eQTLci/download.php)^17^, and were analyzed jointly to infer differential gene expression in cases vs. controls using MetaXcan software with default parameters. The GTEx Elastic-Net Model pipeline (https://github.com/hakyimlab/PredictDBPipeline) was applied to prepare the model used for MetaXcan. The LD references were estimated based on genotypes of European individuals from the 1000 Genome Project. Using FDR < 0.05, the method indentified 5,990 coding genes with significant models for glomerular eQTL, and 5,371 coding genes for tubular eQTL. Significant association was defined as *p*<0.05/2/6050=4.1×10^−6^, i.e., corrected for two tissues and 6,050 genes found in either tubular or glomerular eQTL data.

### Gene-prioritization analysis

Gene prioritization at each of our top loci was performed using two complementary similarity-based gene prioritization approaches (PoPS v0.1^18^ and MAGMA^14^), which integrate GWAS summary statistics with gene set enrichment analysis based on a variety of biological annotation datasets including gene expression, curated pathways, protein-protein interactions, and mouse gene knock-out studies.

For PoPS gene prioritization, MAGMA is first used to calculate gene-level association statistics for 18,383 protein-coding genes in the genome, which is used to assess feature enrichment. PoPS then calculates polygenic priority (PoP) scores for each gene based on its membership to enriched features. For each of our top loci, we annotated the PoPS prioritized gene as the one with the highest PoP score within a 500kb flanking window of each of our lead SNPs.

Of note, the *PRNCR1* gene annotated as the nearest gene to SNP rs185299109 was not included in the PoPS protein-coding gene dataset and the CKD-associated SNP rs185299109 located in an intergenic region was also excluded from this analysis. MAGMA gene prioritization was conducted using a recently developed extension to the method as described and implemented in Benchmarker software^19^, enabling the explicit derivation of gene prioritization results from gene set enrichment analysis. Like the Benchmarker approach, we classified genes as members of each gene set using the top 50, 100, and 200 ranked genes, and obtained similar results from all three. To identify the PoPS features that contributed to the prioritization of *COL4A3*, we limited it to the selected marginal gene features (PoPS step 1), multiplied the *COL4A3* beta hats (PoPS step 2) by the *COL4A3* feature’s scores, and ranked the features by the highest overall score.

### Expression quantitative trait loci (eQTL)

eQTL associations were sought from the eQTLGen database for eQTL in whole blood from >30,000 participants (http://www.eqtlgen.org/)^20^. Kidney specific eQTL associations were queried from eQTL datasets for glomeruli, tubules^17^, and a meta-analysis of four eQTL studies with 451 kidney samples. The meta-analysis of four eQTLs datasets obtained from the Susztak lab, The Cancer Genome Atlas (TCGA), the Genotype-Tissue Expression (GTEx v8), and the Nephrotic Syndrome Study Network (NephQTL)^17,21–23^, was performed using METAL with fixed effects inverse-variance meta-analysis^24^.

### Methylation quantitative trait loci (mQTL)

mQTL associations were sought for the lead SNPs in 188 healthy kidney samples (eGFR > 60 and fibrosis < 10%), with Bonferroni threshold (*p*<1.5×10^−11^) considered genome-wide significant. DNA methylation of CpG sites were profiled in 188 healthy kidney samples by the Infinium MethylationEPIC Kit and BeadChips (Illumina, USA) and were transformed by an inverse-normal transformation after quality control using SeSAMe.^25^ Genotypes for these samples were profiled by from Axiom Tx and Axiom Biobank arrays and imputed using the multiethnic panel reference from 1000Genomes Phase 3 (NCBI build 37, released in October 2014). The association between CpG site and the SNPs within 1Mb were estimated by linear regression model using MatrixQTL,^26^ with covariates including collection site, age, sex, top five genotype PCs, degree of bisulfite conversion, sample plate, and sentrix position and PEER factors. For the significant CpG sites, we then sought for evidence of association between blood methylation levels and eGFR, or eGFR decline, in 500 individuals with diabetes^27^; we furthermore tested association with DKD in our epigenome-wide association study in 1304 UK-ROI and FinnDiane participants, analyzed using the Infinium MethylationEPIC Kit and BeadChips (Illumina, USA), following the QC and analysis procedures described earlier for UK-ROI^28^. Meta-analysis of the two data sets was performed with METAL software^24^, based on *p*-values and direction of effect.

### Human kidney gene expression

For the 29 lead genes or transcripts underlying or located near the lead SNPs, or based on gene-level analyses, TWAS, PoPS, or kidney eQTL data, we studied gene expression in kidneys in human transcriptomics data from nephrectomy samples (433 tubule and 335 glomerulus samples)^29^ and kidney biopsies from the Pima Indian cohort (67 glomerular and 47 tubulointerstitial tissues),^30^ and tested for correlation with relevant pathological phenotypes. The microdissected nephrectomy samples were from individuals with varying degree of diabetic and hypertensive kidney disease, and gene expression was defined with RNA sequencing. Pearson correlation *p*-values below 2.2×10^−4^ were considered significant, corrected for multiple testing for 29 genes, two tissue compartments, and four phenotypes (eGFR, fibrosis, glomerulosclerosis and group comparison). The study was approved by the institutional review board of the University of Pennsylvania.

In the Pima Indian cohort, gene expression profiling in the first biopsy was performed with Affymetrix gene chip arrays^30^, and with Illumina RNA-sequencing for the second biopsy, as described earlier^6^. Available phenotypes included progression to ESRD, measured GFR (mGFR), albumin-to-creatinine ratio (ACR), glycated hemoglobin (HbA1c) and six kidney morphological parameters for both biopsies, and change in the phenotypes between the first and the second study biopsies (27 phenotypes in total).^31^ Pearson correlation *p*-values below 3.2×10^−5^were considered significant, corrected for 29 genes, 2 tissues, and 27 phenotypes; *p*-values below 8.6×10^−4^ (i.e. without correction for 27 phenotypes) were considered suggestive. The study was approved by the Institutional Review Board of the National Institute of Diabetes and Digestive and Kidney Diseases.

### Further annotation of the lead variants

Chromatin 3D conformation interactions with gene transcription start sites (TSS) were queried for the most significant SNPs from the promoter capture Hi-C (PCHiC) data from the www.chicp.org web interface, including data for GM12878 lymphoblastoid cell line and CD34 cells^32^, hESC derived cardiomyocytes^33^, 16 primary blood cells^34^, and pancreatic islets^35^. Interactions with score ≥ 5 were considered significant. We queried chromatin accessibility in kidney single-nucleus ATAC-sequencing (snATACseq) data available at https://susztaklab.com/human_kidney/igv/ (accessed 24 June 2021)^36^. Detailed gene expression in kidney single cell RNA sequencing (scRNAseq) data was queried in the Human Diabetic Kidney data set (23,980 nuclei) by Wilson *et al*.^37^, accessed through http://humphreyslab.com/SingleCell. Further epigenetic annotation was sought from the regulomeDB^38^, and differential renal gene expression in DKD versus healthy controls from the Ju CKD^39^ and Woroniecka^40^ data sets in the NephroSeq portal (www.nephroseq.org). Of note, samples in the queried Woroniecka^40^ data are a subset (N=22) of the more recent RNA sequencing based nephrectomy samples mentioned above (N=433)^29^.

### LD score regression (LDSR) and Mendelian Randomization (MR)

LDSR^41^ was performed at LDhub (http://ldsc.broadinstitute.org/) for 78 glycemic, autoimmune, anthropometric, bone, smoking behavior, lipid, kidney, uric acid, cardiometabolic, and aging related traits, based on the GWAS summary statistics of the ten DKD phenotypes explored. Variants were filtered to those with MAF ≥1%. LDSR associations with *p*<6.4×10^−4^ were defined significant after Bonferroni correction for 78 traits. To identify causal relationships for significant traits in the LDSR against DKD, we performed summary-based two-sample MR implemented in the R package TwoSampleMR^42^. For the SNP-trait associations, we selected genetic variants as instrumental variables (IV) that were independently associated with the selected traits (*p*<5×10^−8^; r^2^ < 0.001 based on the 1000Genomes EUR panel; LD window=10,000 kb) from published GWAS. Palindromic SNPs with intermediate allele frequency (MAF close to 50%) were removed. Traits with less than five IVs were excluded from the MR analysis. Primarily, we used inverse variance-weighted (IVW) regression, but causality was further assessed using methods less sensitive to pleiotropy/heterogeneity (weighted median and MR-Egger regression)^43^. Heterogeneity of SNP estimates in MR was assessed with Cochran’s Q statistic *p*-value and the I^2^ statistic. The MR–Egger intercept test was used to detect unbalanced horizontal pleiotropy.

### Data sharing information

The GWAS meta-analysis results can be accessed via the T1D, T2D, and Cardiometabolic Disease (CMD) Knowledge Portals (https://t1d.hugeamp.org/; https://t2d.hugeamp.org/; https://hugeamp.org/), and downloaded on their respective downloads pages.

## RESULTS

### GWAS meta-analysis

The GWAS meta-analysis of the DNCRI (T1D), and SUMMIT-T1D and SUMMIT-T2D cohorts included up to 26,785 individuals with diabetes from 25 studies; 11,380 individuals with any DKD (micro- or macroalbuminuria or ESRD) and 15,405 individuals with normal AER (**Supplemental Table 2**). QQ plots and genomic correlation λ_GC_ of the meta-analysis indicated no marked inflation of the results; similarly, also LDSR intercepts were close to 1.00 (1.00 – 1.05) for each phenotype, demonstrating no evidence of population stratification bias in the GWAS meta-analysis (**Supplemental Figure 1)**.

The meta-analysis identified a novel association between the combined CKD-DKD phenotype and rs72831309 (minor allele frequency [MAF] =4%, odds ratio [OR] = 2.08, 95% confidence interval [CI] 1.62 - 2.67, *p*=9.8×10^−9^; **Figure 2A**; **Table 1; Supplemental Table 4**); the variant is located in an intron of the *TENM2* gene encoding the teneurin transmembrane protein 2. Exploration of the RegulomeDB revealed that rs72831309 alters a predicted CREB1 transcription factor binding site (**Figure 2C**). Kidney eQTL data indicated that rs72831309 was nominally associated with expression of a *TENM2* antisense transcript *CTB-178M22*.*2* in kidneys (*p*=6.9×10^−3^). Furthermore, chromatin conformation data in the GM12878 cell line indicated that the rs72831309 containing DNA fragment interacts with the *TENM2* transcription start site (TSS), as well as with three antisense transcripts (*CTB-180C19*.*1, CTB-105L4*.*2*, and *CTB-78F1*.*1*) within the *TENM2* gene^32^. Finally, human kidney scRNAseq showed that *TENM2* is expressed in podocytes and in the parietal epithelial cells (**Figure 2D**)^37^.

**Figure 2:**
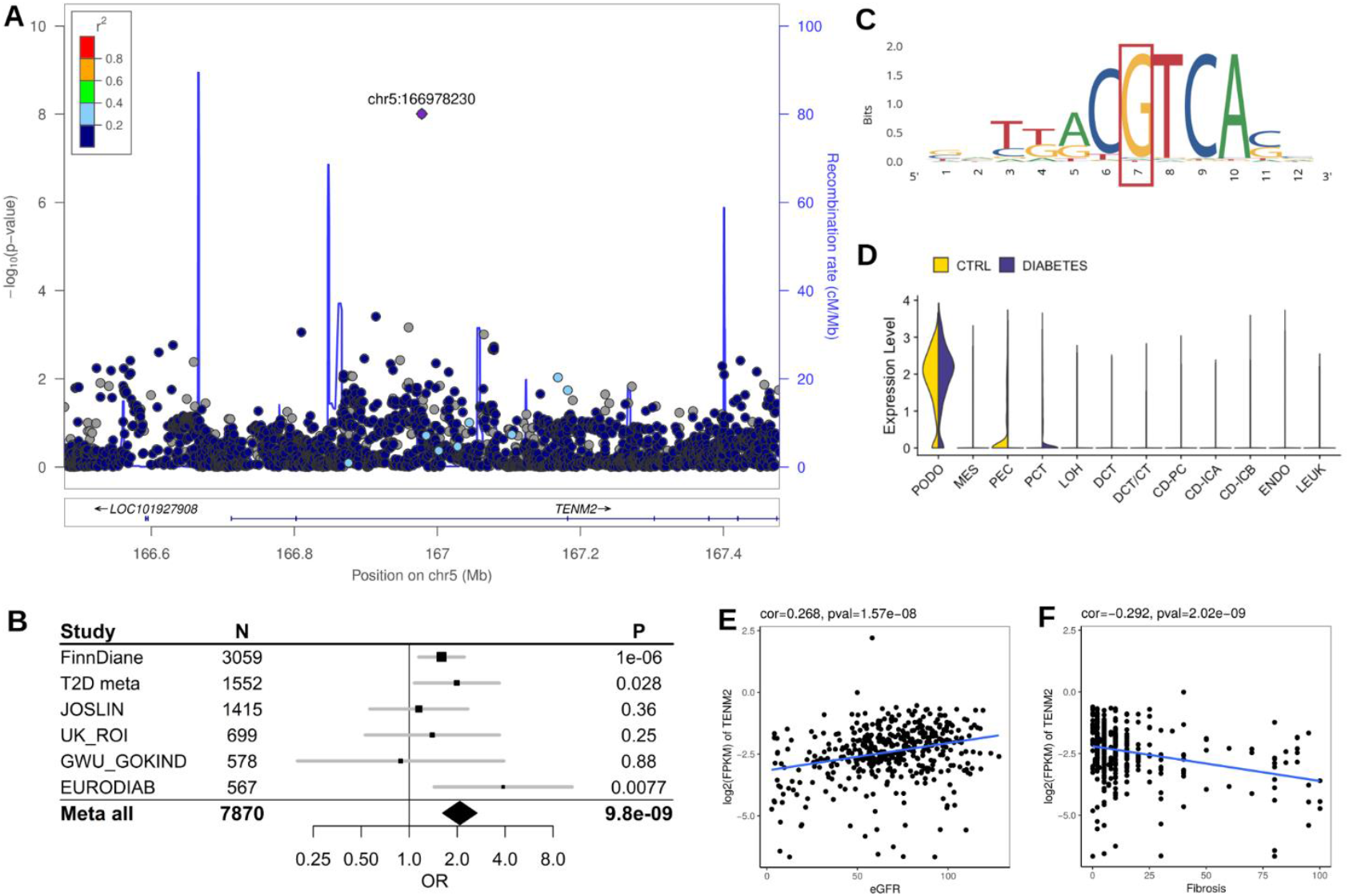
*TENM2* gene rs72831309 is associated with CKD-DKD. **A:** Regional association plot of the meta-analysis results. **B:** Forest plot of association across the contributing cohorts. **C**: SNP rs72831309 overlaps a predicted CREB1 motif sequence; data from RegulomeDB.org. **D:** Human kidney single cell RNA expression of *TENM2* showing strongest expression in podocytes (PODO), parietal epithelial cells (PEC) and proximal convoluted tubules (PCD). **E** and **F**: Tubular TENM2 expression is correlated with higher eGFR (E) and less fibrosis (F).

**Table 1:**
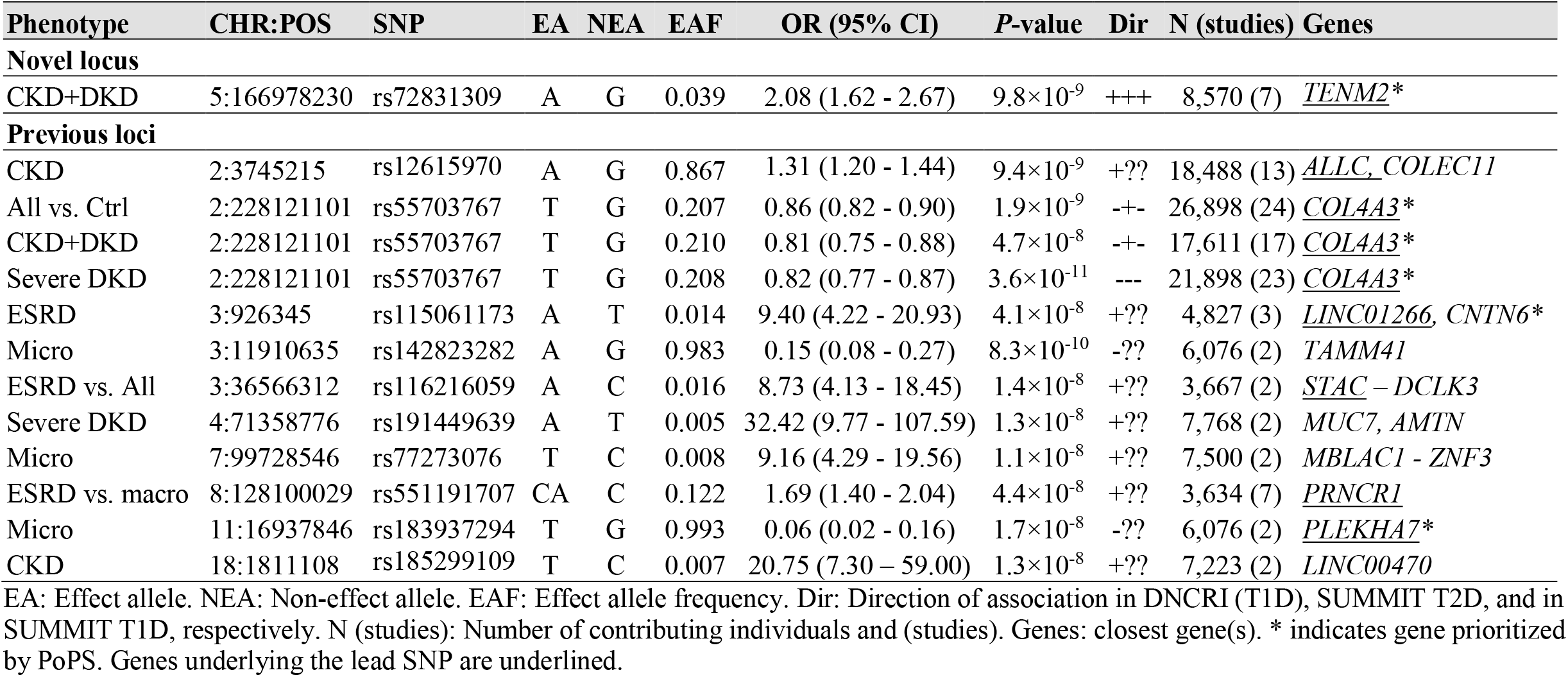
GWAS meta-analysis result summary: loci with *p*<5×10^−8^.

In addition to rs72831309, ten previously identified, mostly low-frequency or rare variants were associated with various kidney phenotypes (**Table 1, Supplemental Figure 2**). Apart from the *COL4A3* locus, none of these variants were found in the SUMMIT cohorts (filtered to MAF≥1%), and thus, these associations represent the originally reported associations from the DNCRI^6^. Here, we identified a secondary signal with a higher frequency (MAF=0.44) in only partial LD (D’=0.51, r^2^=0.08) with the lead signal at the *COL4A3* locus (rs6436688 with *p*=1.8×10^−7^ for Severe DKD; **Supplemental Figure 3**), which after conditional analysis for the rs55703767 lead variant remained nominally significant (*p*=0.002).

Two variants, chr3:141792314:I and rs186434345, were associated with ESRD (N=940, *p*=4.6×10^−10^), and with the CKD-DKD phenotype (N=2,571, *p*=4.0×10^−8^) in the SUMMIT-T2D and SUMMIT-T1D cohorts.The variants were absent (or poor imputation quality, r^2^<0.3) in all DNCRI cohorts derived from the more recent 1000Genomes phase 3 reference. When the original SUMMIT-FinnDiane GWAS was included in the analysis, both associations were non-significant (chr3:141792314:I: *p*=0.056, N=3,207, and rs186434345: *p*=0.002, N=4,782), and thus, excluded from further consideration.

### Gene-level analysis

To improve power and jointly test all available common genetic markers within a gene, SNP summary statistics from the GWAS meta-analysis were aggregated by gene and tested jointly for association using two similar programs, MAGMA and PASCAL. In addition to *COL20A1* and *SNX30* identified in the previously published DNCRI gene-level analysis^6^, we identified eight novel gene associations including *GPR158, LSM14A*, and *MFF* associated with severe DKD; *INIP* associated with any DKD; *PTPRN* and *RESP18* associated with CKD; and *DCLK1* and *EIF4E* associated with ESRD vs. macroalbuminuria (*p*<2.7×10^−6^; **Table 2, Supplemental Figures 4A-J**). Kidney eQTL data for the lead SNPs in the *INIP-SNX30* region suggested *SNX30* as the target gene (rs786959 eQTL *p*=4.6×10^−7^; **Supplemental Table 5**).

**Table 2:** Significant Gene-level DKD association results from MAGMA and PASCAL.

| Phenotype | Gene | Chr | BP Start | BP End | N SNPs | P-value | Method | Genes |
| --- | --- | --- | --- | --- | --- | --- | --- | --- |
| CKD | *PTPRN* | 2 | 220149345 | 220179295 | 72 | $2.13 \times 10^{-6}$ | MAGMA | 18,461 |
| CKD | *RESP18* | 2 | 220187131 | 220202899 | 62 | $2.27 \times 10^{-6}$ | MAGMA | 18,461 |
| Severe DKD | *MFF* | 2 | 228189866 | 228222552 | 439 | $2.07 \times 10^{-6}$ | PASCAL | 21,790 |
| ESRD vs. macro | *EIF4E* | 4 | 99794607 | 99856786 | 150 | $5.79 \times 10^{-7}$ | MAGMA | 18,442 |
| ESRD vs. macro | *EIF4E* | 4 | 99799606 | 99851786 | 269 | $9.28 \times 10^{-7}$ | PASCAL | 21,762 |
| DKD | *INIP* | 9 | 115443786 | 115485387 | 111 | $4.89 \times 10^{-7}$ | MAGMA | 18,475 |
| DKD | *INIP* | 9 | 115448790 | 115480387 | 248 | $1.87 \times 10^{-6}$ | PASCAL | 21,784 |
| DKD | *SNX30* | 9 | 115506911 | 115642267 | 505 | $1.30 \times 10^{-6}$ | MAGMA | 18,475 |
| Severe DKD | *GPR158* | 10 | 25459290 | 25896158 | 1875 | $1.63 \times 10^{-6}$ | MAGMA | 18,467 |
| ESRD vs. macro | *DCLK1* | 13 | 36337789 | 36710514 | 1162 | $1.39 \times 10^{-6}$ | MAGMA | 18,442 |
| Severe DKD | *LSM14A* | 19 | 34658352 | 34725420 | 180 | $1.90 \times 10^{-6}$ | MAGMA | 18,467 |
| CKD extremes | *COL20A1* | 20 | 61919538 | 61967285 | 146 | $1.94 \times 10^{-7}$ | MAGMA | 18,440 |
| ESRD vs. all | *COL20A1* | 20 | 61919538 | 61967285 | 145 | $5.26 \times 10^{-7}$ | MAGMA | 18,439 |

BP Start/End: Basepair position of the start and the end of the gene region. N SNPs: Number of SNPs in the gene region. Genes: number of genes tested.

### Genome-wide integration of GWAS results with kidney eQTL data

We performed TWAS for each of the ten DKD meta-analyses to predict differential gene expression between cases and controls in human glomeruli and tubules, based on eQTL data in glomerular and tubulointerstitial samples from histologically normal kidneys.^17^ Expression levels of *AKIRIN2*, encoding a nuclear protein involved in stimulating pro-inflammatory pathways such as NF-κB^44^, were predicted to be higher in the tubular tissue of cases with severe DKD as compared to controls with normal AER (*p*=1.1×10^−6^, **Supplemental Table 6**). The eQTL data with 39 SNPs explained 5% of the variance in tubular *AKIRIN2* expression (*p*=0.01), which was further correlated with fibrosis (**Figure 3B**).

**Figure 3:**
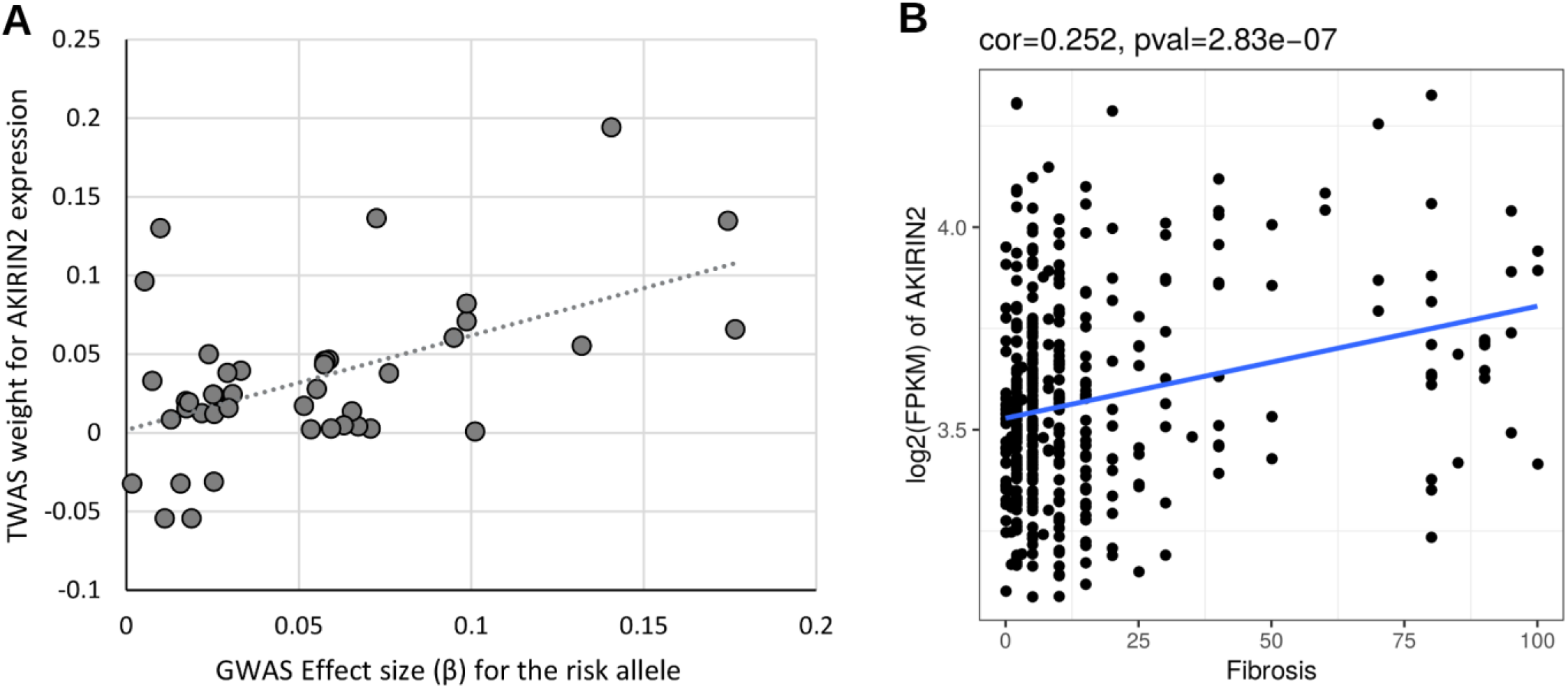
TWAS indicates increased *AKIRIN2* expression in Severe DKD. **A:** The GWAS SNP effect sizes for association with Severe DKD (normal AER vs. macroalbuminuria or ESRD) are correlated with TWAS eQTL weights to predict *AKIRIN2* expression, suggesting that elevated *AKIRIN2* levels in tubules are associated with Severe DKD (*p*=1.1×10^−6^). **B:** *AKIRIN2* expression is correlated with renal fibrosis, correlation = 0.252, *p*-value=2.83×10^−7^.

### Gene prioritization

To identify the underlying causal genes within each of our top loci, we used the PoPS^18^ method that leverages genome-wide enrichment of biological annotations in combination with GWAS summary statistics to prioritize candidate genes. To increase precision, we intersected the results with both the simple nearest gene approach, as well as MAGMA gene prioritization. Four genes were both the PoPS prioritized gene and the nearest gene to the lead associated SNP, including *COL4A3, PLEKHA7, CNTN6*, and *TENM2* (**Table 1**). Of note, the *CNTN6* locus contained only two genes and the *TENM2* locus only one, with a relatively low PoP Score (−0.25). When taking the intersect between PoPS genes and genes that were within MAGMA’s top 10% of prioritized genes genome-wide, *COL4A3* was the only prioritized gene **(Supplemental Figure 5)**.

Given the robust evidence for *COL4A3* as a causal gene for DKD^6^, we interrogated which gene features contributed to its prioritization in MAGMA, reasoning that additional genes that are key members of these same gene features are also candidate genes for DKD and may provide a deeper understanding of disease mechanism. The specific enriched gene set that prioritized *COL4A3* for Severe DKD using the MAGMA gene prioritization approach was the fibulin 2 protein-protein interaction network (“FBLN2 PPI subnetwork”), which together with 26 correlated reconstituted gene-sets makes up the “basement membrane” meta-gene set derived in Marouli *et al*.^45^ These gene-sets prioritized 86 genes in MAGMA, including three other genes specific to the “FBLN2 PPI subnetwork” **(Supplemental Table 7)**.

### Kidney mQTL

To see whether the identified loci might affect the risk of DKD through DNA methylation, kidney mQTL data were queried for the top three variants at each lead locus from the GWAS meta-analyses, gene-level analyses, and TWAS. Altogether, 17 variants were significantly associated with kidney DNA methylation levels at six CpG sites (*p*<1.5×10^−11^**; Supplemental Table 8**). These included SNPs in the *LSM14A* gene, associated with severe DKD and cg14143166 methylation levels (*p*=1.9×10^−28^). Interestingly, cg14143166 methylation in blood was nominally associated with DKD status in our epigenome-wide association study (EWAS) in the UK-ROI and FinnDiane cohorts (*p*=0.03), further supporting the hypothesis that the DKD association at *LSM14A* might be mediated through methylation changes. Similarly, blood methylation levels at significant kidney mQTL CpG sites (rs7664964–cg25974308 *p*=1.1×10^−11^) in *EIF4E* were nominally associated also with the eGFR slope in diabetes (*p*=0.04)^27^.

### Gene expression and pathological phenotypes

Altogether, we identified 29 lead genes or transcripts either underlying or located near the lead SNPs, or based on gene-level analyses, TWAS, PoPS, or kidney eQTL data. Among these, the expression levels of 14 genes were significantly correlated with eGFR, glomerulosclerosis, or fibrosis in transcriptomics data obtained from 433 tubular and 335 glomerular nephrectomy samples with varying degree of diabetic and hypertensive kidney disease (*p*<2.2×10^−4^; **Figure 4, Supplemental Table 9**).^29^ The strongest correlations were observed for the tubular expression of *DCLK1* and *COL4A3 (*positive correlation), and *TENM2, COLEC11, ALLC, PLEKHA7*, and *SNX30* (negative correlation) with the level of fibrosis; and tubular *TENM2, PLEKHA7, ALLC*, and *SNX30* expression positively correlated with eGFR (*p*<1×10^−7^, |*r*| 0.29-0.56). In the Pima Indian kidney biopsy data, tubular *DCLK1* expression levels at the first biopsies were suggestively correlated (*p*<8.6×10^−4^, corrected for 29 genes and two tissues) with higher level of fibrosis, and *LSM14A* negatively correlated with the change in mesangial volume between the two study biopsies (**Figure 4)**; however, these correlations did not remain significant after a conservative correction for altogether 27 correlated tested phenotypes from two different time points. Multiple genes were nominally (*p*<0.05) correlated with these renal parameters (**Supplemental Figure 6, Supplemental Table 10**).

**Figure 4:**
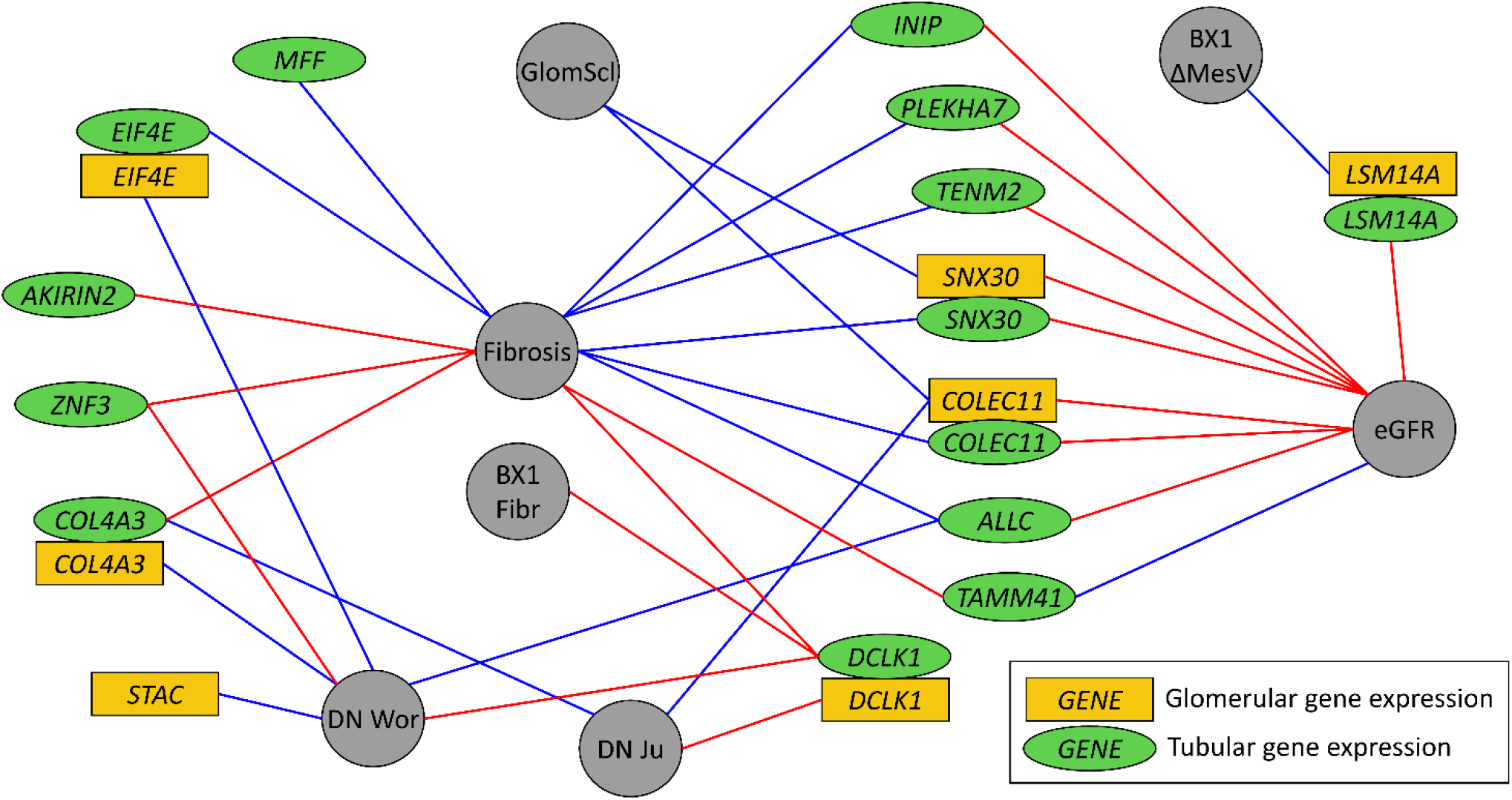
Tubular and glomerular gene expression of the lead genes correlates with multiple morphological and pathological renal parameters and with DKD. Golden rectangles indicate glomerular gene expression, green ellipses tubular gene expression, and grey circles the morphological phenotypes. Blue edges indicate negative correlation, red edges positive correlation. Correlation with fibrosis, glomerulosclerosis (GlomScl), and eGFR were measured in the nephrectomy samples^29^, shown are correlations with p<2.2×10^−4^ (corrected for 29 genes, 2 tissues, and 4 tests). For the biopsy data in Pima Indians, suggestive correlations with p<8.6×10^−4^ are shown (corrected only for 29 genes and 2 tissues), including BX1 Fibr (fibrosis at first biopsy) and BX1 ΔMesV, change in the mesangial volume between the first and the second biopsies. Association with DKD (diabetic nephropathy, DN) was queried in two data sets, DN Wor (Woroniecka et al.^40^) and DN Ju (Ju et al^39^) with p<4.3×10^−4^, or p<0.05 and fold change>1.5.

### Genetic correlation and Mendelian randomization (MR) with related traits

LDSR was used to assess the shared inheritance across the genome between the DKD phenotypes and the related metabolic traits. The analysis revealed significant genetic correlation (*p*<6.4×10^−4^) between DKD and 15 traits including multiple obesity-related traits, mother’s age at death, T2D, coronary artery disease, HDL cholesterol, urate, and two smoking-related traits (**Figure 5**; **Supplemental Figure 7**). Among these 15 traits, all but “mother’s age at death”, which had fewer than five genome-wide significant SNP associations (*p*<5×10^−8^), were used in subsequent MR analysis. MR suggested that the overweight and obesity related traits were causal risk factors for DKD (**Supplemental Table 11, Figure 5B**). The causal effects were directionally consistent across methods, with no evidence of heterogeneity (I^2^=0-42.9%, *p*>0.05, **Supplemental Table 11**) or unbalanced horizontal pleiotropy (**Supplemental Table 12**). The MR Egger method, more robust for pleiotropic effects, further supported a causal role of higher BMI, waist, and hip circumference on DKD risk (*p*<0.05, **Supplemental Table 11, Supplemental Figure 8**). The other significant LDSR associations that indicated no causal association in MR analysis may represent reverse causality or shared upstream effects.

**Figure 5:**
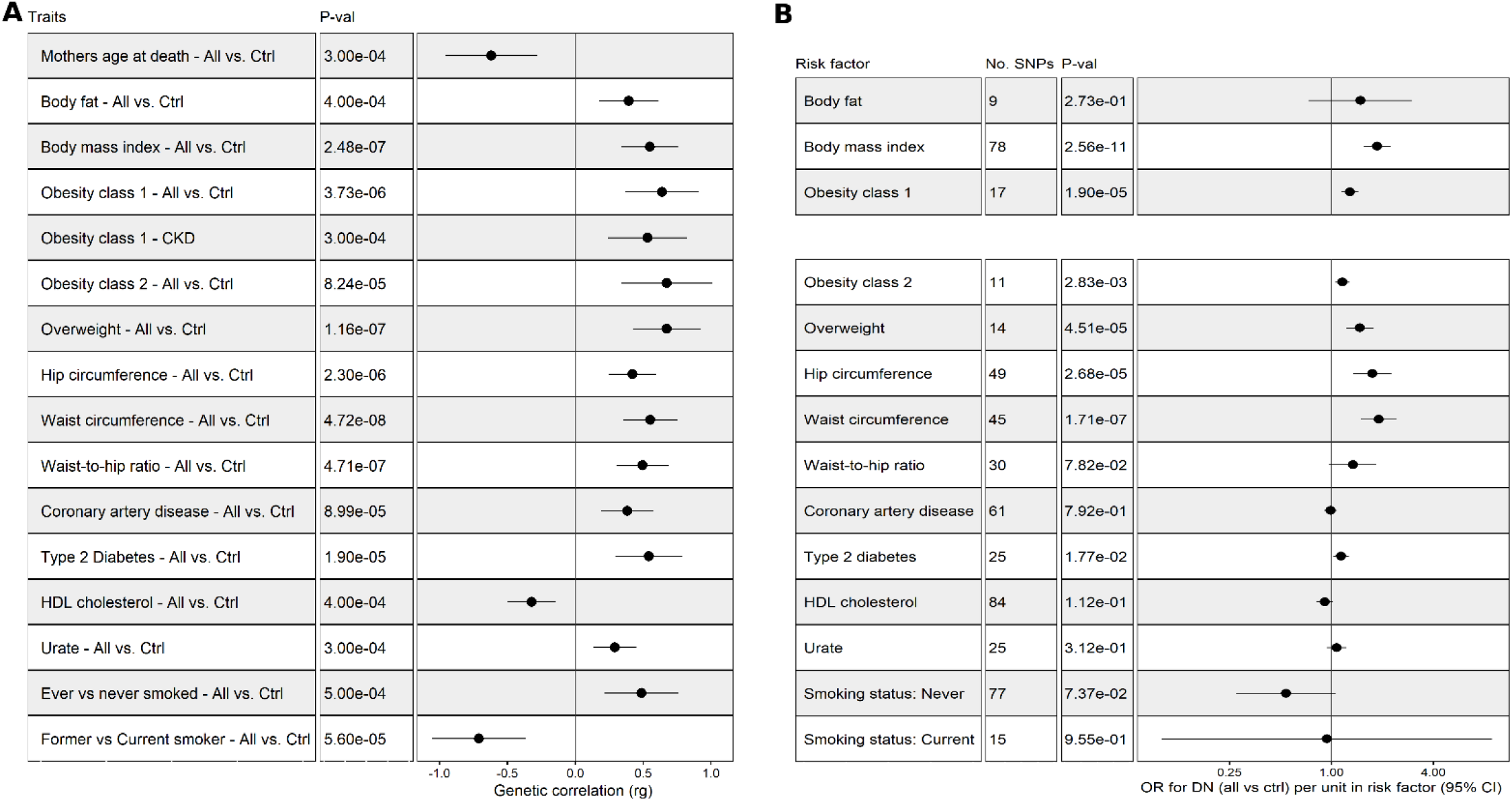
Genetic correlation between DKD phenotypes and various traits based on LDSR (A), and estimates of causal associations based on Mendelian Randomization (B). For LDSR only significant trait combinations are shown (p-value < 0.05/78 = 6.4×10^−4^). **A: B:** Mendelian Randomization results for DKD (All vs. Ctrl) with inverse variance weighted (IVW) method for the traits significant in LDSR, and with at least 5 genome-wide significant SNPs.

## DISCUSSION

We have performed the largest GWAS meta-analysis to date on kidney complications in diabetes, including ten different phenotypic definitions in up to 26,785 individuals with either T1D or T2D, and integrated the results with emerging kidney omics data (**Figure 1**). In the single variant analysis with the combined CKD-DKD phenotype (cases with eGFR<45 ml/min/1.73 m^2^ and microalbuminuria or worse, vs. controls with normal AER and eGFR ≥ 60 ml/min/1.73 m^2^), we identified one novel locus, rs72831309 intronic in the *TENM2* gene. *TENM2* encodes the teneurin transmembrane protein 2, which is involved in cell-cell adhesion. *TENM2* rs72831309 was one of the lead loci associated with DKD in the original SUMMIT T1D GWAS meta-analysis, but it failed to reach genome-wide significance at the time^4^. Supporting the functionality of rs72831309, the SNP alters a predicted CREB1 transcription factor binding site. Even though no direct eQTL evidence was found for rs72831309 affecting *TENM2* expression, it was nominally associated with expression of a *TENM2* antisense transcript *CTB-178M22*.*2* in kidneys (*p*=0.007), while chromatin conformation data in the GM12878 cell line also pointed towards *TENM2* as the effector gene. Whereby kidney scRNAseq indicated *TENM2* expression in podocytes and proximal tubular cells, lower tubular *TENM2* expression was associated with renal fibrosis (*p*=2.0×10^−9^) and lower eGFR (*p*=1.6×10^−8^) in the nephrectomy samples; it was also lower among individuals with DKD versus controls (*p*=6.6×10^−4^, not significant after correction for multiple testing). Other variants in the *TENM2* gene were associated with multiple traits including smoking status (*p*=1.6×10^−17^) and BMI (*p*=2.6×10^−8^) in the UK Biobank. Furthermore, it was previously reported that the DNA methylation status in *TENM2* is among the strongest predictors of incident type 2 diabetes^46^.

Gene level analysis identified ten genes associated with various definitions of DKD. The *DCLK1* gene encodes a doublecortin-like kinase and is a known cancer stem cell marker involved in epithelial to mesenchymal cell transition.^47^ The histone modification based ChromHMM 15-state model for fetal kidney indicated strong transcription overlapping one of the three lead SNPs in the *DCLK1* locus (rs61948262), and ChIP-seq data supported *ZSCAN4* binding to the locus in the HEK293 kidney epithelial cell line. The lead SNPs were kidney mQTLs for *DCLK1* CpG sites (*p*=6.8×10^−22^), further supporting their functional relevance. Furthermore, multiple lines of evidence highlight the importance of *DCLK1* in DKD **(Figure 6, Supplemental Table 10)**: The correlation between tubular *DCLK1* expression and fibrosis was amongst the strongest correlations both in the nephrectomy samples (*p*=7.4×10^−16^) and in the Pima Indian biopsies (*p*=3.0×10^−4^), and glomerular *DCLK1* expression was nominally associated with glomerular width, mesangial volume, and podocyte foot process width in the Pima Indian biopsies (*p*<0.05). Furthermore, both glomerular and tubular *DCLK1* expression were elevated in DKD in two independent data sets (fold change [FC] 1.98, *p*=1.2×10^−4^ for glomeruli;^39^ FC 2.1, *p*=0.003 for tubules^40^). Finally, we have previously identified a subset of transcripts, including *DCLK1*, targeted by the early growth response-1 (egr-1) transcription factor in a murine model of DKD. In this study, *DCLK1* expression was upregulated in diabetic versus non-diabetic ApoE^-/-^ mouse kidneys.^48^ Taken together, these expression data in human and experimental DKD identify *DCLK1* as a novel target.

**Figure 6:**
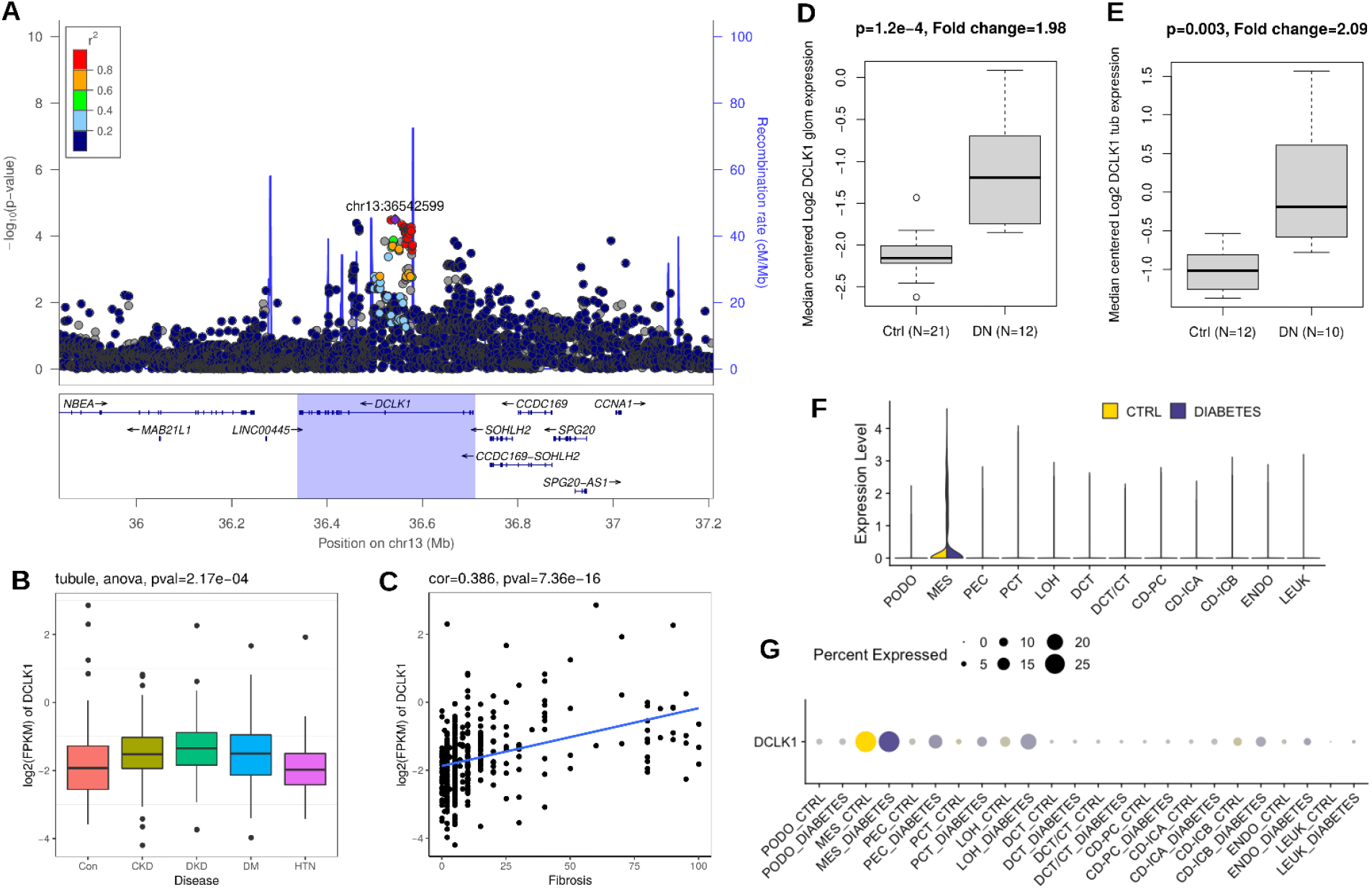
*DCLK1* is associated with ESRD. **A:** The *DCLK1* gene region was associated with ESRD vs. macroalbuminuria in the MAGMA gene-level analysis (p=1.39×10^−6^). **EB and C:** Tubular *DCLK1* expression is highest in DKD **(B)**, and correlated with the level of fibrosis **(C)** in the nephrectomy samples. **D:** Glomerular *DCLK1* expression is higher in DKD than in healthy controls (Ju CKD^39^: Fold change 1.98, p=1.2×10^−4^). **E:** Tubular *DCLK1* expression is higher in DKD than in healthy controls (Woroniecka data set^40^: Fold change 2.1, p=0.003). **F and G:** Kidney *DCLK1* expression is strongest in mesangial cells in human scRNAseq data of individuals with diabetes and healthy controls^37^.

Of note, one of the previously identified associations with ESRD at rs116216059, intronic in the *STAC* gene, is located <200kb downstream of *DCLK3* from the same protein family. The SNP is located on a chromatin accessibility peak border in podocytes (snATACseq peak value 1.1; **Supplemental Figure 2F**). PCHiC data in GM12878 cell line supports both *STAC* and *DCLK3* as the target gene, and the expression levels of both genes were nominally associated with glomerular pathology parameters in the transcriptomics data sets (**Supplemental Table 10**).

The *SNX30* gene, encoding the Sorting Nexin Family Member 30, was associated with DKD in the gene-level analysis. While the association signal spans also the neighboring *INIP* gene, kidney eQTL data for the top SNPs in the locus pointed towards *SNX30*, with the DKD risk associated rs786959 A allele associated with higher *SNX30* expression (*p*=4.6×10^−7^). On the contrary, the nephrectomy sample transcriptomics data indicated that higher tubular *SNX30* was correlated with higher eGFR (*p*=5.8×10^−14^) and lower level of fibrosis (*p*<2×10^−16^); glomerular expression was correlated with less glomerulosclerosis (*p*=8×10^−5^). Finally, kidney *SNX30* expression was associated with higher eGFR in the general population using TWAS based on kidney eQTL^17^ and GWAS on eGFR^49^ (*p*=0.046).

The TWAS analysis based on our GWAS results, integrated with microdissected tubular and glomerular eQTL data, predicted that the *AKIRIN2* gene expression is elevated in tubules in individuals with severe DKD compared with individuals with normal AER. Furthermore, the *AKIRIN2* gene expression was highly correlated with the level of fibrosis (*p*=2.8×10^−7^). *AKIRIN2* encodes the akirin-2 protein, a conserved nuclear factor required for the innate immune response. Akirin-2 is a downstream effector of the toll-like receptor, tumor necrosis factor and IL-1 beta signaling pathways. It binds to nuclear NF-κB complexes and is required for the transcription of a subset of NF-κB–dependent genes such as *IL-6, CXCL10*, and *CCL5;*^50^ NF-κB activation drives inflammatory responses and is activated in DKD.^51^

The strongest regulatory evidence in RegulomeDB was obtained for rs1260634 intronic in the *LSM14A* gene: rs1260634 is located in a *ZNF362* binding sequence in HEK293 cell line, exerts strong transcription in 125 tissue types including fetal kidney chromatin state model, and affects a predicted transcription factor binding motif for KLF4, KLF12, and SP8 (**Supplemental Figure 9)**. Furthermore, in our kidney mQTL data, rs1260634 showed strong association (*p*=2.1×10^−28^) with a CpG site cg14143166, where methylation in blood was associated with DKD in our EWAS data (*p*=0.03). Tubular *LSM14A* expression correlated with higher eGFR (*p*=2.9×10^−6^), and glomerular expression with the decrease in mesangial volume (6.5×10^−4^). *LSM14A* encodes a Sm-like protein, thought to participate in pre-mRNA splicing, and implicated in innate antiviral responses^52^. Other noteworthy novel genes include *EIF4E* and *PTPRN*: *EIF4E* encodes a common mRNA translation initiation factor; its activation and/or suppression are influenced by mTOR signaling cascades involved in DKD^53^ as well as high glucose and high insulin environments in renal epithelial cells.^54^ *PTPRN* (protein tyrosine phosphatase receptor type N) encodes IA-2, a major T1D autoantigen involved in glucose-stimulated insulin secretion^55^. In mice, IA-2 is required to maintain normal levels of renin expression in kidneys^56^. Finally, the *MFF* gene encoding the mitochondrial fission factor, which was identified in our gene level analysis, has been previously related to DKD^57,58^. However, the association may be driven by the neighboring *COL4A3* associations, as was first suggested in the DNCRI GWAS eQTL analysis.^6^

In the transcriptomics analyses, as expected, all 13 significant correlations with the level of fibrosis were observed for tubular gene expression, whereby the two observed correlations for glomerulosclerosis were for glomerular expression of *SNX30* and *COLEC11* (**Figure 3**). Interestingly, eight out of ten correlations with eGFR were obtained for tubular rather than glomerular gene expression, supporting the importance of tubular damage for the loss of renal function.

Previous genetic risk scores and LDSR analyses have supported a role of T2D genetic factors, obesity and smoking for the development of DKD^4^. While there is a strong epidemiological link between DKD and coronary artery disease in diabetes^1^, this is the first study to report also a genetic correlation between these major diabetic micro- and macrovascular complications. Among the lipid traits, significant correlation with DKD was found only for lower HDL cholesterol, despite previous MR studies within general population cohorts implicating HDL as a marker of dyslipidemia rather than a causal factor^59^. Indeed, our subsequent MR found no evidence of causality between HDL cholesterol and DKD; in concordance with our previous MR on BMI,^60^ only body weight related measurements were causal risk factors for DKD. Of note, our current MR was in line with our previous MR in T1D suggesting that serum urate levels are not a causal risk factor for DKD^61^; similar negative results were also reported for non-diabetic CKD^62^.

The majority of kidney disease in individuals with T1D is considered to occur due to diabetic nephropathy, histologically characterized by thickening of glomerular basement membrane and mesangial expansion, as well as renal tubular, interstitial and arteriolar lesions.^63^ In individuals with T2D, only a proportion of DKD is purely due to diabetic nephropathy, whereas aging, obesity, and hypertension also contribute to the development of kidney complications. Thus, including individuals with T2D in the meta-analysis increases the heterogeneity of the underlying disease, and may select findings for those directly related to hyperglycemia as the common endpoint shared by both forms of diabetes. However, as T2D represents 95% of all diabetes cases, including those individuals greatly increases statistical power for our current work and future GWAS meta-analyses integrating multiple subtypes of diabetes to identify shared genetic risk factors for DKD.

## Author contributions

NS and JC performed the main analyses and drafted the manuscript. VN, XS, HL, EA, NvZ, EHD, DF, LS performed statistical analyses, contributed to interpretation of the results, and revised the manuscript. RS, CF, EV, VH, EB, GM, DA, RD, HCL, RGN, AJM, CP, CG, APM, LG, MIM, MK, KS, JNH, JCF, PHG contributed to the data acquisition and interpretation of the results, and critically revised the manuscript.

## Supplemental Table of contents

Supplemental Table 1: A total of ten case – control definitions for diabetic kidney disease (DKD; used as a general term to describe renal complications in diabetes)

Supplemental Table 2: Number of individuals in each cohort for each phenotypic comparison

Supplemental Table 3: Key characteristics of the genotyping and statistical analyses in DNCRI and SUMMIT cohorts.

Supplemental Table 4: Association details for the novel genome-wide significant locus rs72831309 in *TENM2*.

Supplemental Table 5: Kidney eQTL associations with p<1×10^−4^ in tubular or glomerular eQTL data, or in the kidney eQTL meta-analysis for the lead SNPs.

Supplemental Table 6: TWAS results with p<1×10^−4^.

Supplemental Table 7: Highly correlated reconstituted gene-sets that make-up the “basement membrane” meta-gene set derived in Marouli et al.

Supplemental Table 8: Significant kidney mQTL associations (p<1.46×10^−11^ Bonferroni-adjusted genome-wide significance; 1×10^−7^ suggestive significance) for lead loci.

Supplemental Table 9: Correlation between glomerular and tubular gene expression and glomerulosclerosis, fibrosis, and eGFR in nephrectomy samples.

Supplemental Table 10: Gene centric summary of the lead genes

Supplemental Table 11: Mendelian Randomization (MR) results for DKD (All vs Ctrl)

Supplemental Table 12. Egger intercepts for Mendelian Randomization analyses on DKD (all vs. Ctrl phenotype).

Supplemental Figure 1: Manhattan and QQ-plots of the ten DKD GWAS meta-analysis results.

Supplemental Figure 2: Regional association plots for the GWAS meta-analysis lead loci (A-K).

Supplemental Figure 3: Regional association plot for the *COL4A3* gene region associated with Severe DKD, indicating a secondary association peak at chr2:228259302 (rs6436688, effect allele (A) frequency 56%, OR = 1.13 (95% confidence interval 1.08 – 1.19), p-value 1.79×10^−7^).

Supplemental Figure 4: Regional association plots for the gene-level analysis results from MAGMA and PASCAL analysis.

Supplemental Figure 5: Gene prioritization for the *COL4A3* gene at lead SNP rs5570367 associated with Severe DKD using multiple intersecting gene prioritization approaches (PoPS, nearest gene, and MAGMA).

Supplemental Figure 6: Tubular and glomerular gene expression of the lead genes correlate with multiple morphological and pathological renal parameters. Golden

Supplemental Figure 7: Genetic correlation between DKD and related traits based on LD score regression.

Supplemental Figure 8: Mendelian Randomization scatter plots for SNP effects for the metabolic traits vs. DKD (All vs. Ctrl).

Supplemental Figure 9: rs1260634 intronic in the *ALLC* gene affects the predicted binding motifs for KLF12, KLF4, and SP8 (top to bottom).

