## Supplemental material for "Genome-wide meta-analysis and omics integration identifies novel genes associated with diabetic kidney disease"

**Supplemental Table 1: A total of ten case – control definitions for diabetic kidney disease (DKD; used as a general term to describe renal complications in diabetes)**

| Phenotype | Cases | Controls | Note |
|---|---|---|---|
| **All vs. Ctrl** | microalbuminuria or macroalbuminuria or ESRD | Normal AER | Phenotype abbreviated as "DN" (diabetic nephropathy) in SUMMIT |
| **Severe DKD** | macroalbuminuria or ESRD | Normal AER | Phenotype abbreviated as "MACRO" in SUMMIT; as "DN" in DNCRI |
| **Micro** | microalbuminuria | Normal AER | |
| **Macro** | macroalbuminuria | Normal AER | Not analyzed in SUMMIT |
| **ESRD** | ESRD | Normal AER | |
| **ESRD vs. All** | ESRD | no ESRD | |
| **ESRD vs. macro** | ESRD | macroalbuminuria | Not analyzed in SUMMIT |
| **CKD** | eGFR < 60 ml/min/1.73m2 | eGFR ≥ 60 ml/min/1.73m2 | |
| **CKD extremes** | ESRD or eGFR < 15 ml/min/1.73m2 | eGFR ≥ 60 ml/min/1.73m2 | Not analyzed in SUMMIT |
| **CKD-DKD** | ESRD, or eGFR < 60 ml/min/1.73m2 AND microalbuminuria or macroalbuminuria | normal AER and eGFR ≥ 60 ml/min/1.73m2 | |

Normal AER: AER <30mg/24h, or equivalent
Microalbuminuria: 30 mg/24h ≤AER < 300 mg/24h, or equivalent
Macroalbuminuria: AER ≥ 300 mg/24h, or equivalent
ESRD: End-stage renal disease, dialysis or renal transplant (or eGFR< 15 ml/min/1.73m2 in SUMMIT)

#### Supplemental Table 2: Number of individuals in each cohort for each phenotypic comparison

| Study | Severe DKD | | | Macro | | | ESRD | | | ESRD vs. All | | | ESRD vs. Macro | | | All vs. Ctrl | | | Micro | | | CKD | | | CKD extremes | | | CKD-DKD | | |
|---|---|---|---|---|---|---|---|---|---|---|---|---|---|---|---|---|---|---|---|---|---|---|---|---|---|---|---|---|---|---|
| **DNCRI** | **Cases** | **Ctrls** | **Total** | **Cases** | **Ctrls** | **Total** | **Cases** | **Ctrls** | **Total** | **Cases** | **Ctrls** | **Total** | **Cases** | **Ctrls** | **Total** | **Cases** | **Ctrls** | **Total** | **Cases** | **Ctrls** | **Total** | **Cases** | **Ctrls** | **Total** | **Cases** | **Ctrls** | **Total** | **Cases** | **Ctrls** | **Total** |
| Austria | 6 | 71 | **77** | 4 | 71 | **75** | 2 | 71 | **73** | 2 | 88 | **90** | 2 | 4 | **6** | 19 | 71 | **90** | 13 | 71 | **84** | 11 | 80 | **91** | 2 | 80 | **82** | 6 | 66 | **72** |
| CACTI | 35 | 422 | **457** | 29 | 422 | **451** | 6 | 422 | **428** | 6 | 503 | **509** | 6 | 29 | **35** | 87 | 422 | **509** | 52 | 422 | **474** | 45 | 477 | **522** | 6 | 477 | **483** | 24 | 407 | **431** |
| EDIC | 134 | 193 | **327** | 75 | 193 | **268** | 59 | 193 | **252** | 59 | 359 | **418** | 59 | 75 | **134** | 225 | 193 | **418** | 91 | 193 | **284** | 130 | 288 | **418** | 59 | 288 | **347** | 89 | 180 | **269** |
| EDC | 84 | 1016 | **1100** | 61 | 1016 | **1077** | 23 | 1016 | **1039** | 23 | 1282 | **1305** | 23 | 61 | **84** | 289 | 1016 | **1305** | 205 | 1016 | **1221** | 80 | 1218 | **1298** | 23 | 1218 | **1241** | 47 | 999 | **1046** |
| FinnDiane | 1371 | 2240 | **3611** | 535 | 2268 | **2803** | 854 | 2265 | **3119** | 854 | 3418 | **4272** | 854 | 517 | **1371** | 2069 | 2202 | **4271** | 719 | 2257 | **2976** | 1226 | 3038 | **4264** | 838 | 3069 | **3907** | 993 | 2066 | **3059** |
| FRANCE | 332 | 627 | **959** | 181 | 625 | **806** | 151 | 627 | **778** | 151 | 920 | **1071** | 151 | 181 | **332** | 448 | 627 | **1075** | 124 | 625 | **749** | 281 | 740 | **1021** | 159 | 740 | **899** | 225 | 578 | **803** |
| GWU GOKIND | 290 | 311 | **601** | 29 | 311 | **340** | 261 | 311 | **572** | 261 | 340 | **601** | 261 | 29 | **290** | 290 | 311 | **601** | 0 | 311 | **311** | 273 | 325 | **598** | 261 | 325 | **586** | 269 | 309 | **578** |
| ITALY | 180 | 161 | **341** | 38 | 161 | **199** | 142 | 161 | **303** | 142 | 201 | **343** | 142 | 38 | **180** | 180 | 163 | **343** | 0 | 161 | **161** | 157 | 172 | **329** | 148 | 175 | **323** | 155 | 154 | **309** |
| JOSLIN | 719 | 1082 | **1801** | 475 | 1082 | **1557** | 244 | 1082 | **1326** | 244 | 2027 | **2271** | 244 | 475 | **719** | 1189 | 1082 | **2271** | 470 | 1082 | **1552** | 533 | 1574 | **2107** | 262 | 1695 | **1957** | 402 | 1013 | **1415** |
| LatDiane | 25 | 80 | **105** | 18 | 80 | **98** | 7 | 80 | **87** | 7 | 131 | **138** | 7 | 18 | **25** | 58 | 80 | **138** | 33 | 80 | **113** | 16 | 109 | **125** | 7 | 109 | **116** | 9 | 78 | **87** |
| LitDiane | 19 | 39 | **58** | 9 | 39 | **48** | 10 | 39 | **49** | 10 | 69 | **79** | 10 | 9 | **19** | 40 | 39 | **79** | 21 | 39 | **60** | 21 | 50 | **71** | 10 | 50 | **60** | 16 | 36 | **52** |
| RomDiane | 98 | 89 | **187** | 70 | 89 | **159** | 28 | 89 | **117** | 28 | 207 | **235** | 28 | 70 | **98** | 146 | 89 | **235** | 48 | 89 | **137** | 53 | 167 | **220** | 28 | 167 | **195** | 39 | 87 | **126** |
| Scotland | 195 | 3962 | **4157** | 144 | 3984 | **4128** | 57 | 3962 | **4019** | 57 | 4632 | **4689** | 57 | 138 | **195** | 727 | 3962 | **4689** | 540 | 3984 | **4524** | 404 | 4712 | **5116** | 80 | 4712 | **4792** | 90 | 4450 | **4540** |
| STENO | 488 | 414 | **902** | 469 | 414 | **883** | 19 | 414 | **433** | 19 | 897 | **916** | 19 | 470 | **489** | 489 | 427 | **916** | 0 | 414 | **414** | 200 | 690 | **890** | 28 | 690 | **718** | 106 | 398 | **504** |
| SWEDEN | 51 | 346 | **397** | 35 | 346 | **381** | 20 | 346 | **366** | 20 | 497 | **517** | 20 | 32 | **52** | 51 | 346 | **397** | 85 | 346 | **431** | 42 | 287 | **329** | 20 | 287 | **307** | 21 | 252 | **273** |
| UK_ROI | 704 | 730 | **1434** | 466 | 730 | **1196** | 200 | 730 | **930** | 200 | 1196 | **1396** | 200 | 466 | **666** | 704 | 730 | **1434** | 0 | 730 | **730** | 587 | 513 | **1100** | 200 | 513 | **713** | 266 | 433 | **699** |
| WESDR | 217 | 293 | **510** | 113 | 293 | **406** | 104 | 293 | **397** | 104 | 452 | **556** | 104 | 113 | **217** | 263 | 293 | **556** | 46 | 293 | **339** | 207 | 398 | **605** | 104 | 398 | **502** | 140 | 260 | **400** |
| **Total DNCRI** | **4,948** | **12,076** | **17,024** | **2,751** | **12,124** | **14,875** | **2,187** | **12,101** | **14,288** | **2,187** | **17,219** | **19,406** | **2,187** | **2,725** | **4,912** | **7,274** | **12,053** | **19,327** | **2,447** | **12,113** | **14,560** | **4,266** | **14,838** | **19,104** | **2,235** | **14,993** | **17,228** | **2,897** | **11,766** | **14,663** |
| **SUMMIT T1D** | **Cases** | **Ctrls** | **Total** | **Cases** | **Ctrls** | **Total** | **Cases** | **Ctrls** | **Total** | **Cases** | **Ctrls** | **Total** | **Cases** | **Ctrls** | **Total** | **Cases** | **Ctrls** | **Total** | **Cases** | **Ctrls** | **Total** | **Cases** | **Ctrls** | **Total** | **Cases** | **Ctrls** | **Total** | **Cases** | **Ctrls** | **Total** |
| Eurodiab | 203 | 491 | **694** | | | | 84 | 491 | **575** | 84 | 705 | **789** | | | | 298 | 491 | **789** | 95 | 491 | **586** | 113 | 467 | **580** | | | | 210 | 357 | **567** |
| NFS-ORPS | 47 | 199 | **246** | | | | | | | | | | | | | 197 | 199 | **396** | 150 | 199 | **349** | | | | | | | | | |
| SDR | 168 | 292 | **460** | 85 | 277 | **362** | 75 | 294 | **369** | 75 | 529 | **604** | 57 | 85 | **142** | 266 | 290 | **556** | 98 | 290 | **388** | 163 | 365 | **528** | 57 | 349 | **406** | 118 | 239 | **357** |
| **Total SUMMIT T1D** | **418** | **982** | **1,400** | **85** | **277** | **362** | **159** | **785** | **944** | **159** | **1,234** | **1,393** | **57** | **85** | **142** | **761** | **980** | **1,741** | **343** | **980** | **1,323** | **276** | **832** | **1,108** | **57** | **349** | **406** | **328** | **596** | **924** |
| **SUMMIT T2D** | **Cases** | **Ctrls** | **Total** | **Cases** | **Ctrls** | **Total** | **Cases** | **Ctrls** | **Total** | **Cases** | **Ctrls** | **Total** | **Cases** | **Ctrls** | **Total** | **Cases** | **Ctrls** | **Total** | **Cases** | **Ctrls** | **Total** | **Cases** | **Ctrls** | **Total** | **Cases** | **Ctrls** | **Total** | **Cases** | **Ctrls** | **Total** |
| GoDARTS Affy | 218 | 816 | **1,034** | 138 | 816 | **954** | 80 | 816 | **896** | 48 | 1,491 | **1,539** | 80 | 138 | **218** | 885 | 816 | **1,701** | 667 | 816 | **1,483** | 1,025 | 1,553 | **2,578** | 80 | 1,101 | **1181** | 168 | 716 | **884** |
| GoDARTS Illumina | 179 | 680 | **859** | 130 | 675 | **805** | 48 | 680 | **728** | 80 | 1,621 | **1,701** | 48 | 130 | **178** | 859 | 680 | **1,539** | 680 | 680 | **1,360** | 972 | 513 | **1,485** | 48 | 1092 | **1140** | 120 | 587 | **707** |
| MNI | 66 | 165 | **231** | | | | | | | | | | | | | 188 | 165 | **353** | 122 | 162 | **284** | | | | | | | | | |
| SDR | 713 | 580 | **1,292** | 424 | 556 | **980** | 243 | 580 | **823** | 243 | 1,359 | **1,602** | 268 | 424 | **692** | 1,250 | 580 | **1,830** | 520 | 580 | **1,100** | 997 | 666 | **1,663** | 240 | 628 | **868** | 609 | 307 | **916** |
| Steno | 163 | 131 | **294** | | | | | | | | | | | | | 163 | 131 | **294** | | | | 100 | 174 | **274** | | | | | | |
| **Total SUMMIT T2D** | **1,339** | **2,372** | **3,710** | **692** | **2,047** | **2,739** | **371** | **2,076** | **2,447** | **371** | **4,471** | **4,842** | **396** | **692** | **1,088** | **3,345** | **2,372** | **5,717** | **1,989** | **2,238** | **4,227** | **3,094** | **2,906** | **6,000** | **368** | **2,821** | **3,189** | **897** | **1,610** | **2,507** |
| **Total ALL** | **6,705** | **15,430** | **22,134** | **3,528** | **14,448** | **17,976** | **2,717** | **14,962** | **17,679** | **2,717** | **22,924** | **25,641** | **2,640** | **3,502** | **6,142** | **11,380** | **15,405** | **26,785** | **4,779** | **15,331** | **20,110** | **7,636** | **18,576** | **26,212** | **2,660** | **18,163** | **20,823** | **4,122** | **13,972** | **18,094** |

Number of samples in the SUMMIT T1D and T2D cohorts represents those individuals included in the original analysis, containing related individuals; The current meta-analysis was based on effect size estimates from SUMMIT which were derived after excluding related individuals (see Supplemental Table 3 analysis method).

**Supplemental Table 3: Key characteristics of the genotyping and statistical analyses in DNCRI and SUMMIT cohorts.**

| | DNCRI | SUMMIT T1D and T2D |
|---|---|---|
| N studies | 17 | 3 (T1D), 5 (T2D) |
| N samples (max) | 19,406 | 1,741 (T1D), 6,000 (T2D) |
| Genotyping chips | HumanCoreExome Bead arrays 12-1.0, 12-1.1, and 24-1.0 | Illumina Omni express array, Affymetrix SNP 6.0 array, Illumina 610Quad assay |
| Imputation reference panel | 1000Genomes Phase 3 | 1000Genomes Phase 1 |
| Imputation software | Minimac3/Minimac3-omp (version 1.0.14) | Prephasing with SHAPE-IT v2; Imputation with IMPUTEv2 |
| Covariates | Age, sex, diabetes duration, genetic principal components, study specific covariates (e.g. site or genotyping batch) | Age, gender, duration of diabetes, genetic principal components |
| Main post-analysis SNP QC filters | •Imputation quality score: INFO ≥ 0.3<br>•Minor allele count ≥ 10 in cases and in controls<br>•Marker must be present in at least 2 studies | •Imputation quality score: INFO ≥ 0.4<br>•Minor allele count ≥ 10 in cases and in controls<br>•MAF ≥ 0.01<br>•T2D: Marker must be present in at least 2 studies (not applied for the three additional phenotypes available only in SDR, GoDARTS 1 and 2) |
| Analysis method and software | SNPtest, additive score test | SNPtest, additive score test. Note: in original studies, *P* values of association were estimated using EMMAX mixed model including related individuals, while only effect size estimates were obtained from SNPtest (excluding related individuals). |
| Meta-analysis | Inverse-variance fixed effects meta-analysis (METAL software) | Inverse-variance fixed effects meta-analysis (GWAMA or METAL) |

**Supplemental Table 4: Association details for the novel genome-wide significant locus rs72831309 in TENM2.** In the meta-analysis, rs72831309 (chr5:166978230) minor A allele was considered the effect allele, major G as the non-effect allele.

| COHORT | EAF | BETA | SE | OR | OR_L95 | OR_U95 | P | INFO | N |
|---|---|---|---|---|---|---|---|---|---|
| GWU_GOKIND | 0.02 | -0.11 | 0.76 | 0.88 | 0.20 | 3.93 | 0.88 | 0.38 | 578 |
| UK_ROI | 0.03 | 0.57 | 0.49 | 1.40 | 0.53 | 3.68 | 0.25 | 0.53 | 699 |
| JOSLIN | 0.03 | 0.33 | 0.36 | 1.15 | 0.57 | 2.33 | 0.36 | 0.51 | 1415 |
| FinnDiane | 0.05 | 0.82 | 0.17 | 1.60 | 1.15 | 2.22 | $1.0\times10^{-6}$ | 0.66 | 3059 |
| EURODIAB | 0.03 | 1.36 | 0.51 | 3.89 | 1.43 | 10.54 | 0.008 | 0.52 | 567 |
| T2D meta | 0.03 | 0.69 | 0.31 | 1.99 | 1.08 | 3.67 | 0.028 | NA | 1552 |
| **Meta all** | **0.04** | **0.73** | **0.13** | **2.08** | 1.62 | 2.67 | **$9.82\times10^{-9}$** | | **8322** |

EAF: effect allele (minor A allele) frequency. BETA: effect size estimate. SE: standard error for BETA. OR: odds ratio. OR_L95 and OR_U95: Lower and upper confidence intervals. P: p-value. INFO: imputation quality info metrics. N: Number of samples in the study.

**Supplemental Table 5: Kidney eQTL associations with $p<1\times10^{-4}$ in tubular or glomerular eQTL data, or in the kidney eQTL meta-analysis for the lead SNPs.** Three top SNPs were queried for each lead locus from GWAS meta-analysis, gene-level (MAGMA or PASCAL) analysis, or TWAS locus.

| Tissue | SNP | CHR:POS | P eQTL | eQTL GENE | Index gene | P GWAS |
|---|---|---|---|---|---|---|
| Glomerular | rs28577966 | 4:99796005 | 2.13E-07 | *ADH4* | *EIF4E* | 1.05E-06 |
| Glomerular | rs7664964 | 4:99796439 | 2.13E-07 | *ADH4* | *EIF4E* | 8.92E-07 |
| Glomerular | rs11725932 | 4:99799310 | 2.13E-07 | *ADH4* | *EIF4E* | 9.97E-07 |
| Tubular | rs59113552 | 6:88236233 | 5.19E-05 | *SMIM8* | *AKIRIN2* | 2.85E-05 |
| Kidney meta | rs786959 | 9:115429626 | 4.59E-07 | *SNX30* | *INIP* | 9.913E-07 |
| Kidney meta | rs6011746 | 20:61964452 | 5.75E-05 | *CHRNA4* | *COL20A1* | 1.90E-06 |

Tissue: kidney eQTL meta-analysis, or glomerular or tubule compartment-specific expression.

**Supplemental Table 6: TWAS results with $p<1\times10^{-4}$.** P-values < $4.1\times10^{-6}$ are significant after correction for multiple testing.

| | | | | TWAS association | | | Prediction performance | | | | |
|---|---|---|---|---|---|---|---|---|---|---|---|
| **Tissue** | **Pheno** | **gene** | **gene_name** | **Effect** | **p-value** | **var_g** | **r2** | **p-value** | **q-value** | **n_snps_used** | **n_snps_in_model** |
| tub | Severe DKD | ENSG00000135334.8 | *AKIRIN2* | 0.308 | ***1.11E-06*** | 0.092 | 0.05 | 0.013 | 0.012 | 39 | 42 |
| tub | Macro | ENSG00000135334.8 | *AKIRIN2* | 0.383 | ***1.70E-06*** | 0.092 | 0.05 | 0.013 | 0.012 | 39 | 42 |
| glom | Micro | ENSG00000268208.1 | *AC008372.1* | -7.489 | 1.59E-05 | 0.000 | 0.03 | 0.083 | 0.044 | 1 | 1 |
| glom | Macro | ENSG00000228696.4 | *ARL17B* | -0.195 | 2.06E-05 | 0.331 | 0.51 | 1.28E-19 | 1.77E-18 | 54 | 65 |
| glom | Micro | ENSG00000138028.10 | *CGREF1* | -0.228 | 2.15E-05 | 0.131 | 0.14 | 2.08E-05 | 3.07E-05 | 25 | 26 |
| glom | CKD | ENSG00000078804.8 | *TP53INP2* | 0.499 | 2.73E-05 | 0.068 | 0.03 | 0.080 | 0.042 | 34 | 38 |
| glom | Macro | ENSG00000227057.3 | *WDR46* | -0.583 | 4.05E-05 | 0.024 | 0.03 | 0.085 | 0.044 | 10 | 10 |
| tub | Severe DKD | ENSG00000205269.4 | *TMEM170B* | 0.192 | 4.62E-05 | 0.133 | 0.06 | 0.007 | 0.007 | 90 | 95 |
| tub | CKD | ENSG00000188283.7 | *ZNF383* | 0.327 | 4.72E-05 | 0.046 | 0.05 | 0.012 | 0.011 | 15 | 16 |
| tub | CKD | ENSG00000075413.13 | *MARK3* | -0.258 | 5.05E-05 | 0.085 | 0.05 | 0.013 | 0.012 | 102 | 104 |
| tub | Macro | ENSG00000162836.7 | *ACP6* | -0.146 | 6.44E-05 | 0.377 | 0.47 | 4.19E-18 | 7.16E-17 | 30 | 31 |

Tissue: tub(ular) or glom(erular). TWAS association: Effect: association effect size for the gene. P-value: p-value for the TWAS association. var_g: variance of the gene expression. Prediction performance r2, p-value and q-value: statistics for tissue model's correlation to gene's measured transcriptome; n_snps_used: number of SNPs from GWAS that were used in the analysis; n_snps_in_model: number of SNPs in the model (i.e. in the transcriptomics data)

**Supplemental Table 7: Highly correlated reconstituted gene-sets that make-up the "basement membrane" meta-gene set derived in Marouli et al.** The "Genes prioritized" column contains all genes prioritized in MAGMA by one of these 26 gene-sets, including the "FBLN2 PPI subnetwork" gene set that prioritized *COL4A3* for Severe DKD.

| GENE-SET ID | GENE-SET DESCRIPTION | GENES PRIORITIZED |
|---|---|---|
| MP:0003044 | impaired basement membrane formation | |
| **ENSG00000163520** | **FBLN2 PPI subnetwork** | *ADAMTS15, CD248,* ***COL4A3****, NID2* |
| MP:0004272 | abnormal basement membrane morphology | |
| GO:0043256 | laminin complex | |
| ENSG00000130702 | LAMA5 PPI subnetwork | |
| ENSG00000132561 | MATN2 PPI subnetwork | |
| ENSG00000091136 | LAMB1 PPI subnetwork | |
| ENSG00000116962 | NID1 PPI subnetwork | |
| ENSG00000135862 | LAMC1 PPI subnetwork | |
| ENSG00000168487 | BMP1 PPI subnetwork | |
| GO:0034446 | substrate adhesion-dependent cell spreading | |
| ENSG00000125810 | CD93 PPI subnetwork | |
| ENSG00000134871 | COL4A2 PPI subnetwork | *ABCC9, CCDC102B, COL18A1, CSPG4, CTHRC1, FN1, IGFBP3, LAMC1, LOXL1, OLFML2B, TGFBI* |
| ENSG00000188153 | COL4A5 PPI subnetwork | *ABCA9, ADAMTS5, AEBP1, ART3, BICC1, C3, C7, COL15A1, COL1A1, COL1A2, COL3A1,* ***COL4A3****, COL5A2, COL6A2, ENSG00000259134, ENSG00000259284, FBLN5, FBN1, FIBIN, FNDC1, FSTL1, GALNTL4, GRB14, IGFBP6, LAMA2, LAMB1, LINC00312, LOX, MMP2, NOV, OLFML1, POSTN, SCN7A, SERPING1, SLIT3, SPARC, THBS2, VGLL3, WDR72* |
| ENSG00000112773 | FAM46A PPI subnetwork | |
| ENSG00000100985 | MMP9 PPI subnetwork | |
| ENSG00000110492 | MDK PPI subnetwork | |
| ENSG00000101680 | LAMA1 PPI subnetwork | |
| GO:0043236 | laminin binding | |
| ENSG00000187498 | COL4A1 PPI subnetwork | *ACTA2, COL14A1, COL4A2, CTHRC1, ENPEP, LHFP, LOXL2, TGFBI* |
| GO:0005605 | basal lamina | |
| ENSG00000213949 | ITGA1 PPI subnetwork | |
| ENSG00000114270 | COL7A1 PPI subnetwork | |
| GO:0050840 | extracellular matrix binding | |
| ENSG00000081052 | COL4A4 PPI subnetwork | *ABCC9, ACTA2, ADAMTS1, ADAMTS5, ASPN, C1R, C1S, C7, CCDC80, COL4A1, COL5A1, COL6A1, DCN, EFEMP1, FBLN1, FKBP7, IGFBP3, LGALS1, LOX, LUM, MGP, NID2, PID1, PXDN, SCN7A, SERPINF1, SPARCL1, VCAN* |
| GO:0005604 | basement membrane | *APLNR, COL12A1, CTGF, HTRA1, ITGB4, ITGB6, MCAM, PRSS23* |

**Supplemental Table 8: Significant kidney mQTL associations ($p<1.46\times10^{-11}$ Bonferroni-adjusted genome-wide significance; $1\times10^{-7}$ suggestive significance) for lead loci.** Three most significant SNPs were queried at each associated loci from single variant, gene-level, and transcriptome-wide association study (TWAS).

| Chr | RSID | SNP_Pos | CpG | CpG_Start | P mQTL | Gene | P DKD/eGFR |
|---|---|---|---|---|---|---|---|
| 19 | rs668933 | 34704936 | cg14143166 | 34716204 | 1.94E-28 | *LSM14A* | 0.03 (DKD) |
| 19 | rs1260634 | 34701331 | cg14143166 | 34716204 | 2.09E-28 | *LSM14A* | 0.03 (DKD) |
| 19 | rs535440 | 34694581 | cg14143166 | 34716204 | 2.12E-28 | *LSM14A* | 0.03 (DKD) |
| 13 | rs12428319 | 36542599 | cg21746263 | 36562319 | 6.81E-22 | *DCLK1* | |
| 13 | rs61948262 | 36533891 | cg21746263 | 36562319 | 4.19E-21 | *DCLK1* | |
| 20 | rs117255010 | 61953366 | cg20706388 | 61958549 | 1.79E-12 | *COL20A1* | |
| 20 | rs6011746 | 61964452 | cg20706388 | 61958549 | 2.14E-12 | *COL20A1* | |
| 20 | rs4809528 | 61943661 | cg20706388 | 61958549 | 2.55E-12 | *COL20A1* | |
| 20 | rs74397198 | 61950071 | cg20706388 | 61958549 | 2.88E-12 | *COL20A1* | |
| 20 | rs143391037 | 61971717 | cg20706388 | 61958549 | 4.76E-12 | *COL20A1* | |
| 2 | rs6436131 | 220151858 | cg06895971 | 220147671 | 9.20E-12 | *PTPRN* | |
| 4 | rs7664964 | 99796439 | cg25974308 | 99852386 | 1.10E-11 | *EIF4E* | 0.041 (eGFR slope) |
| 4 | rs28577966 | 99796005 | cg25974308 | 99852386 | 1.10E-11 | *EIF4E* | 0.041 (eGFR slope) |
| 4 | rs11725932 | 99799310 | cg25974308 | 99852386 | 1.10E-11 | *EIF4E* | 0.041 (eGFR slope) |
| 6 | rs34472900 | 88405040 | cg00551398 | 88298473 | 1.12E-11 | *AKIRIN2* | |
| 6 | rs151077971 | 88405605 | cg00551398 | 88298473 | 1.13E-11 | *AKIRIN2* | |
| 6 | rs59113552 | 88236233 | cg00551398 | 88298473 | 1.58E-11 | *AKIRIN2* | |
| 9 | rs786975 | 115451231 | cg13293976 | 115516494 | 2.20E-11 | *INIP* | 0.012 (eGFR slope) |
| 2 | rs4674377 | 220201272 | cg14891200 | 220197663 | 2.72E-10 | *RESP18* | |
| 2 | rs2090163 | 220178435 | cg14891200 | 220197663 | 1.06E-09 | *PTPRN* | |
| 6 | rs59113552 | 88236233 | cg10313604 | 88493367 | 3.18E-09 | *AKIRIN2* | |
| 6 | rs151077971 | 88405605 | cg10313604 | 88493367 | 7.06E-09 | *AKIRIN2* | |
| 6 | rs34472900 | 88405040 | cg10313604 | 88493367 | 7.11E-09 | *AKIRIN2* | |
| 6 | rs151077971 | 88405605 | cg05834092 | 87792915 | 7.82E-09 | *AKIRIN2* | |
| 6 | rs34472900 | 88405040 | cg05834092 | 87792915 | 7.83E-09 | *AKIRIN2* | |
| 19 | rs535440 | 34694581 | cg21245903 | 34711622 | 9.97E-09 | *LSM14A* | |
| 19 | rs1260634 | 34701331 | cg21245903 | 34711622 | 1.00E-08 | *LSM14A* | |
| 19 | rs668933 | 34704936 | cg21245903 | 34711622 | 1.04E-08 | *LSM14A* | |
| 6 | rs59113552 | 88236233 | cg05834092 | 87792915 | 1.63E-08 | *AKIRIN2* | |
| 6 | rs151077971 | 88405605 | cg15059496 | 88185750 | 3.76E-08 | *AKIRIN2* | |
| 6 | rs34472900 | 88405040 | cg15059496 | 88185750 | 3.76E-08 | *AKIRIN2* | |
| 6 | rs59113552 | 88236233 | cg20648632 | 88182160 | 4.22E-08 | *AKIRIN2* | |
| 2 | rs2090163 | 220178435 | cg19020434 | 220199207 | 6.40E-08 | *PTPRN* | |
| 2 | rs4674377 | 220201272 | cg06895971 | 220147671 | 6.77E-08 | *RESP18* | |
| 6 | rs59113552 | 88236233 | cg15059496 | 88185750 | 7.55E-08 | *AKIRIN2* | |
| 2 | rs4674377 | 220201272 | cg19020434 | 220199207 | 7.84E-08 | *RESP18* | |
| 19 | rs535440 | 34694581 | cg01663383 | 34676533 | 8.98E-08 | *LSM14A* | |
| 19 | rs1260634 | 34701331 | cg01663383 | 34676533 | 9.01E-08 | *LSM14A* | |
| 19 | rs668933 | 34704936 | cg01663383 | 34676533 | 9.24E-08 | *LSM14A* | |

Gene: CpG site annotated gene. P DKD/eGFR: P-value for association between blood methylation at the CpG site and DKD (UK-ROI+FinnDiane EWAS) or with eGFR slope (CRIC EWAS).

**Supplemental Table 9: Correlation between glomerular and tubular gene expression and glomerulosclerosis, fibrosis, and eGFR in nephrectomy samples.** All associations with p<0.05 are shown, significant associations ($p<2.2\times10^{-4}$, corrected for 29 genes × 2 tissue compartments × 4 phenotypes) are in bold.

| | | eGFR | | Glomerulosclerosis | | Fibrosis | | Group comparison | |
|---|---|---|---|---|---|---|---|---|---|
| **Gene** | **Tissue** | **r** | **p** | **r** | **p** | **r** | **p** | **Direction** | **p** |
| *ALLC* | glom | | | | | | | | |
| *ALLC* | tub | 0.26 | ***6.9E-08*** | | | -0.50 | ***2.0E-16*** | lowest in DKD | ***8.3E-05*** |
| *COLEC11* | glom | 0.24 | ***1.0E-05*** | -0.27 | ***1.8E-06*** | | | lowest in DKD | ***1.8E-05*** |
| *COLEC11* | tub | 0.21 | ***1.0E-05*** | | | -0.47 | ***2.0E-16*** | lowest in DKD | 1.37E-03 |
| *PLEKHA7* | glom | 0.14 | 9.5E-03 | | | | | lowest in DKD | ***4.3E-06*** |
| *PLEKHA7* | tub | 0.30 | ***1.3E-10*** | | | -0.49 | ***2.0E-16*** | lowest in DKD | ***1.3E-05*** |
| *SNX30* | glom | 0.24 | ***1.2E-05*** | -0.22 | ***8.0E-05*** | | | lowest for DKD | ***5.5E-05*** |
| *SNX30* | tub | 0.35 | ***5.8E-14*** | | | -0.56 | ***2.0E-16*** | | ***1.2E-06*** |
| *DCLK1* | glom | | | | | | | | |
| *DCLK1* | tub | -0.15 | 1.48E-03 | | | 0.39 | ***7.4E-16*** | Highest in DKD | ***2.17E-04*** |
| *TENM2* | glom | 0.13 | 0.02 | -0.18 | 1.7E-03 | | | | |
| *TENM2* | tub | 0.27 | ***1.6E-08*** | | | -0.29 | ***2.0E-09*** | lowest in DKD | 6.6E-04 |
| *COL4A3* | glom | 0.11 | 0.05 | -0.16 | 4.8E-03 | | | | |
| *COL4A3* | tub | | | | | 0.29 | ***3.2E-09*** | | |
| *ZNF3* | glom | | | 0.12 | 0.04 | | | | |
| *ZNF3* | tub | -0.13 | 7.26E-03 | | | 0.26 | ***1.4E-07*** | | |
| *TAMM41* | glom | -0.11 | 0.04 | 0.16 | 4.1E-03 | | | | |
| *TAMM41* | tub | -0.20 | ***2.0E-05*** | | | 0.26 | ***1.5E-07*** | highest in DKD | 5.6E-03 |
| *AKIRIN2* | glom | | | | | | | | |
| *AKIRIN2* | tub | | | | | 0.25 | ***2.8E-07*** | | |
| *EIF4E* | glom | -0.12 | 0.03 | | | | | highest in DKD | ***2.4E-06*** |
| *EIF4E* | tub | | | | | -0.18 | ***1.9E-04*** | | |
| *LSM14A* | glom | | | | | | | | |
| *LSM14A* | tub | 0.22 | ***2.9E-06*** | | | -0.13 | 0.01 | | |
| *INIP* | glom | | | -0.15 | 9.3E-03 | | | | |
| *INIP* | tub | 0.22 | ***5.5E-06*** | | | -0.21 | ***2.2E-05*** | | |
| *MFF* | glom | | | | | | | | |
| *MFF* | tub | | | | | -0.21 | ***2.1E-05*** | | |
| *MBLAC1* | glom | | | -0.11 | 0.05 | | | lowest in DKD | 2.8E-03 |
| *MBLAC1* | tub | | | | | -0.17 | 5.2E-04 | | |
| *STAC* | glom | | | -0.17 | 3.0E-03 | | | | |
| *STAC* | tub | | | | | | | | |

r: Pearson correlation coefficient between the phenotype and log2(Fragments per kilobases of transcript per 1 million mapped reads [FPKM]) of gene expression in glomeruli/tubules. p: p-value. Group comparison: ANOVA test for group comparison (Controls, chronic kidney disease [CKD], diabetic kidney disease [DKD], diabetes mellitus [DM] (without DKD), hypertension [HTN]) vs. log2(FPKM) gene expression.

### Supplemental Table 10: Gene centric summary of the lead genes

| GENE | Pheno | Indication | PoPS | kidney eQTL P min | kidney mQTL P min | lead SNP PCHiC Max score | | Glom/Tub eQTL P min | Nephrectomy: eGFR r | eGFR p | Glomerulosclerosis r | Glomerulosclerosis p | fibrosis r | fibrosis p | Group comparison direction | Group comparison p | NephroSeq DN vs. healthy: Woroniecka DN vs healthy P | Woroniecka FC | Ju DN vs. healthy P | Ju FC | Pima BX1 correlations | Pima BX2 correlations |
|---|---|---|---|---|---|---|---|---|---|---|---|---|---|---|---|---|---|---|---|---|---|---|
| ***TENM2*** | CKD+DKD | Underlying lead SNP | ***Yes*** | | | ***8.61*** | ***glom*** | | 0.1 | 0.022 | -0.2 | 0.002 | | | | | | | | | | |
| | | | | | | | ***tub*** | | 0.3 | ***1.6E-8*** | | | -0.3 | ***2.0E-9*** | lowest in DKD | 6.6E-4 | | | | | | |
| ***DCLK1*** | ESRD vs. macro | Gene-based test | | | ***6.81E-22*** | | ***glom*** | | | | | | | | | | | | **1.2E-4** | **1.98** | ACR: r=0.29 p=0.016; MesVol: r=0.25 p=0.041; | GlomWidth: r=0.32 p=0.024; MesVol: r=0.29 p=0.038; FootProcW: r=0.71 p=0.0067; |
| | | | | | | | ***tub*** | 0.035 | -0.2 | 0.001 | | | 0.4 | ***7.4E-16*** | Highest in DKD | ***2.17E-4*** | 0.003 | 2.09 | 0.001 | 1.32 | ACR: r=0.39 p=0.0069; **Fibrosis: r=0.52 p=0.0003;** | ACR: r=0.28 p=0.04; Fibrosis: r=0.36 p=0.015; |
| ***COL4A3*** | DKD | Missense variant | ***Yes*** | | | ***8.89*** | ***glom*** | | 0.1 | 0.047 | -0.2 | 0.005 | | | | | **8.3E-5** | -1.48 | 0.032 | -0.43 | SV: r=-0.28 p=0.024; | |
| | | | | | | | ***tub*** | | | | | | 0.3 | ***3.2E-09*** | | | | | **2.1E-4** | -1.33 | | |
| ***COLEC11*** | CKD | Nearby | | | | ***9.67*** | ***glom*** | | 0.2 | ***1.0E-5*** | -0.3 | ***1.8E-6*** | | | lowest in DKD | ***1.8E-5*** | | | 0.019 | **-1.59** | | |
| | | | | | | | ***tub*** | | 0.2 | ***1.0E-5*** | | | -0.5 | ***2.0E-16*** | lowest in DKD | 0.001 | | | 0.036 | -1.47 | | ACR: r=-0.32 p=0.016; HbA1c: r=-0.29 p=0.034; |
| ***ALLC*** | CKD | Underlying lead SNP | | | | ***10.42*** | ***glom*** | | | | | | | | | | | | 0.04 | -1.19 | GlomWidth: r=0.24 p=0.054; ΔSV: r=-0.46 p=0.01; HbA1c: r=0.3 p=0.041; | MesVol: r=-0.29 p=0.04; |
| | | | | | | | ***tub*** | | 0.3 | ***6.9E-8*** | | | -0.5 | ***2.0E-16*** | lowest in DKD | ***8.3E-5*** | **3.1E-4** | -1.43 | | | | |
| ***PLEKHA7*** | Micro | Underlying lead SNP | ***Yes*** | | | ***9.49*** | ***glom*** | | 0.1 | 9.5E-3 | | | | | lowest in DKD | ***4.3E-6*** | | | | | | |
| | | | | | | | ***tub*** | | 0.3 | ***1.3E-10*** | | | -0.5 | ***2.0E-16*** | lowest in DKD | ***1.3E-5*** | | | | | | |
| ***SNX30*** | DKD | Gene-based test; kidney eQTL | | **4.6E-7** | | | ***glom*** | 0.012 | 0.2 | ***1.2E-05*** | -0.2 | ***8.0E-5*** | | | lowest in DKD | ***5.5E-5*** | | | | | | |
| | | | | | | | ***tub*** | 0.002 | 0.4 | ***5.8E-14*** | | | -0.6 | ***2.0E-16*** | lowest in DKD | ***1.2E-6*** | | | | | | eGFR: r=0.33 p=0.013; |
| ***AKIRIN2*** | Severe DKD | TWAS | | 0.017 | ***1.12E-11*** | | ***glom*** | | | | | | | | | | | | | | | |
| | | | | | | | ***tub*** | 0.001 | | | | | 0.3 | ***2.8E-7*** | | | | | | | | |
| ***EIF4E*** | ESRD vs. macro | Gene-based test | | | ***1.10E-11*** | | ***glom*** | | -0.1 | 0.027 | | | | | highest in DKD | ***2.4E-6*** | 0.010 | -1.88 | 0.027 | -1.27 | ACR: r=0.25 p=0.041; | |
| | | | | | | | ***tub*** | | | | | | -0.2 | ***1.9E-4*** | | | | | | | | eGFR: r=-0.4 p=0.0028; |
| ***MFF*** | Severe DKD | Gene-based test | | 0.032 | | ***10.19*** | ***glom*** | 0.007 | | | | | | | | | 5.4E-4 | -1.40 | | | ΔMesVol: r=-0.46 p=0.011; ΔeGFR: r=0.43 p=0.017; Progr to ESRD: p=0.007; | |
| | | | | | | | ***tub*** | 0.017 | | | | | -0.2 | ***2.1E-5*** | | | | | | | | |
| ***ZNF3*** | Micro | Underlying tag SNP | | | | | ***glom*** | | | | 0.1 | 0.038 | | | | | | | | | GlomVol: r=0.25 p=0.05; HbA1c: r=-0.25 p=0.043; | |
| | | | | | | | ***tub*** | | -0.1 | 0.007 | | | 0.3 | ***1.4E-7*** | | | **1.1E-4** | -1.33 | | | | |
| ***TAMM41*** | Micro | Nearby | | | | ***10.65*** | ***glom*** | | -0.1 | 0.037 | 0.2 | 0.004 | | | | | | | | | | |
| | | | | | | | ***tub*** | | -0.2 | ***2.0E-5*** | | | 0.3 | ***1.5E-7*** | highest in DKD | 0.006 | | | | | | ACR: r=0.34 p=0.011; |
| ***LSM14A*** | Severe DKD | Gene-based test | | | ***1.9E-28*** | | ***glom*** | 0.004 | | | | | | | | | 7.4E-4 | -1.27 | 0.001 | -1.27 | **ΔMesVol: r=-0.59 p=0.00065;** ΔSV: r=0.5 p=0.0052; | |
| | | | | | | | ***tub*** | 0.017 | 0.2 | ***2.9E-6*** | | | -0.1 | 0.011 | | | | | | | | |
| ***STAC*** | ESRD vs. All | Underlying lead SNP | | | | ***10.87*** | ***glom*** | | | | -0.2 | 0.003 | | | | | 0.030 | -1.53 | 0.013 | -1.38 | | |
| | | | | | | | ***tub*** | | | | | | | | | | 0.018 | -1.18 | | | | |
| ***PTPRN*** | CKD | Gene-based test | | 0.007 | ***9.2E-12*** | | ***glom*** | | | | | | | | | | 0.016 | 1.14 | 0.002 | -1.16 | FootProcW: r=0.29 p=0.018; | |

| | | | | | | | | | | | | | | | |
|---|---|---|---|---|---|---|---|---|---|---|---|---|---|---|---|
| | | | | | | | ***tub*** | | | | | | 0.011 -1.21 9.5E-4 -1.11 | HbA1c: r=0.28 p=0.059; | |
| ***INIP*** | DKD | Gene-based test | | | ***2.2E-11*** | | ***glom*** | 0.004 | | -0.1 0.009 | | | | | |
| | | | | | | | ***tub*** | | 0.2 ***5.5E-6*** | | -0.2 ***2.2E-5*** | | | | |
| ***CNTN6*** | ESRD | PoPS | ***Yes*** | 0.046 | | | ***glom*** | | | | | | | | |
| | | | | | | | ***tub*** | | | | | | | | |
| ***MBLAC1*** | Micro | Nearby | | | | ***TSS*** | ***glom*** | | | -0.1 0.054 | | lowest in DKD 0.003 | | | |
| | | | | | | | ***tub*** | | | | -0.2 5.2E-4 | | | | |
| ***COL20A1*** | CKD extremes | Gene-based test | | | ***1.8E-12*** | | ***glom*** | | | | | | | | FootProcW: r=-0.68 p=0.01; |
| | | | | | | | ***tub*** | 0.018 | | | | | | | |
| ***DCLK3*** | ESRD vs. All | Nearby | | | | ***8.8*** | ***glom*** | | | | | | | | GlomWidth: r=0.33 p=0.021;<br>MesVol: r=0.34 p=0.015;<br>GlomVol: r=0.42 p=0.0072;<br>SV: r=-0.28 p=0.05; |
| | | | | | | | ***tub*** | | | | | | | | |
| ***MUC7*** | Severe DKD | Nearby | | | | ***10.53*** | ***glom*** | | | | | | | | |
| | | | | | | | ***tub*** | | | | | | | | |
| ***RESP18*** | CKD | Gene-based test | | | ***2.7E-10*** | | ***glom*** | | | | | | | | |
| | | | | | | | ***tub*** | | | | | | | | |
| ***AMTN*** | Severe DKD | Nearby | | | | | ***glom*** | | | | | | | | |
| | | | | | | | ***tub*** | | | | | | | | |
| ***GPR158*** | Severe DKD | Gene-based test | | | | | ***glom*** | | | | | | | | |
| | | | | | | | ***tub*** | | | | | | | | |
| ***LINC01266*** | ESRD | Underlying lead SNP | | | | | ***glom*** | | | | | | | | |
| | | | | | | | ***tub*** | | | | | | | | |
| ***PRNCR1*** | ESRD vs. macro | Underlying lead SNP | | | | | ***glom*** | | | | | | | | FootProcW: r=0.62 p=0.023; |
| | | | | | | | ***tub*** | | | | | | | | |
| ***CTB-178M22.2*** | CKD+DKD | lead SNP kidney eQTL | | 0.007 | ***NA*** | | ***glom*** | | ***NA*** | ***NA*** | ***NA*** | ***NA*** | | | |
| | | | | | | | ***tub*** | | ***NA*** | ***NA*** | ***NA*** | ***NA*** | | | |
| ***ADH4*** | ESRD vs. macro | glom eQTL for EIF4E | | 0.008 | ***NA*** | | ***glom*** | **2.1E-7** | ***NA*** | ***NA*** | ***NA*** | ***NA*** | | | |
| | | | | | | | ***tub*** | 0.002 | ***NA*** | ***NA*** | ***NA*** | ***NA*** | | | ACR: r=0.39 p=0.003; |
| ***SMIM8*** | Severe DKD | tub eQTL for AKIRIN2 | | 1.1E-4 | ***NA*** | | ***glom*** | | ***NA*** | ***NA*** | ***NA*** | ***NA*** | | | |
| | | | | | | | ***tub*** | **5.2E-5** | ***NA*** | ***NA*** | ***NA*** | ***NA*** | | | |
| ***CHRNA4*** | CKD extremes | kidney eQTL for COL20A1 | | **5.8E-5** | ***NA*** | | ***glom*** | 0.009 | ***NA*** | ***NA*** | ***NA*** | ***NA*** | 0.030 1.17 | FootProcW: r=0.27 p=0.029; | GlomWidth: r=-0.29 p=0.041; |
| | | | | | | | ***tub*** | 0.007 | ***NA*** | ***NA*** | ***NA*** | ***NA*** | | | eGFR: r=0.31 p=0.024; |

Indication: why gene was listed as a lead gene. PoPS: was prioritized by PoPS? Kidney eQTL: minimum P-value for eQTL association between the lead SNPs and the gene in the kidney eQTL meta-analysis. Kidney mQTL: minimum P-value for kidney methylation, between the lead SNPs and the gene, as assigned in the mQTL annotation. Lead SNP PCHiC: highest score for chromatin 3D conformation capture data at the chicp.org for the GWAS meta-analysis lead loci. Glom/Tub eQTL: minimum P-value for eQTL association between the lead SNPs and the gene in glomerular/ tubular eQTL data. Nephrectomy Gene expression vs. phenotype correlations: Pearson correlation (r) and p-value for correlation between glomerular/tubular gene expression and the phenotype. NephroSeq DN vs. healthy: Fold change (FC) and p-value for differential glomerular/tubular gene expression in DN vs. healthy in the Woroniecka and Ju CKD data sets. Pima BX1/BX2 correlations: Pearson correlation coefficient r and p-value for glomerular/tubular gene expression vs. morphological parameters at the first (BX1) or second (BX2) biopsy.

**Supplemental Table 11: Mendelian Randomization (MR) results for DKD (All vs Ctrl)**

| Exposure | method | nsnp | OR (95%CI) | se | p-val | Q-pval | $I^2$ (%) |
|---|---|---|---|---|---|---|---|
| **Body fat** | Inverse variance weighted | 9 | 1.48 (0.73-2.98) | 0.36 | 0.27 | 0.02 | 60.2 |
| **Body fat** | Weighted median | 9 | 1.26 (0.64-2.49) | 0.35 | 0.50 | NA | NA |
| **Body fat** | MR Egger | 9 | 1.04 (0.01-98.21) | 2.32 | 0.99 | 0.01 | 65.1 |
| **Body mass index** | Inverse variance weighted | 78 | 1.86 (1.55-2.23) | 0.09 | 2.56e-11 | 0.50 | 0.00 |
| **Body mass index** | Weighted median | 78 | 1.76 (1.31-2.36) | 0.15 | 1.85e-04 | NA | NA |
| **Body mass index** | MR Egger | 78 | 1.98 (1.28-3.08) | 0.22 | 3.18e-03 | 0.48 | 0.36 |
| **Obesity class 1** | Inverse variance weighted | 17 | 1.28 (1.14-1.44) | 0.06 | 1.90e-05 | 0.10 | 31.6 |
| **Obesity class 1** | Weighted median | 17 | 1.24 (1.08-1.42) | 0.07 | 2.26e-03 | NA | NA |
| **Obesity class 1** | MR Egger | 17 | 1.21 (0.87-1.69) | 0.17 | 0.28 | 0.08 | 35.2 |
| **Obesity class 2** | Inverse variance weighted | 11 | 1.16 (1.05-1.28) | 0.05 | 2.83e-03 | 0.10 | 37.2 |
| **Obesity class 2** | Weighted median | 11 | 1.13 (1.02-1.26) | 0.05 | 0.02 | NA | NA |
| **Obesity class 2** | MR Egger | 11 | 1.21 (0.89-1.65) | 0.16 | 0.25 | 0.07 | 42.9 |
| **Overweight** | Inverse variance weighted | 14 | 1.47 (1.22-1.77) | 0.09 | 4.51e-05 | 0.07 | 38.6 |
| **Overweight** | Weighted median | 14 | 1.31 (1.05-1.64) | 0.11 | 0.02 | NA | NA |
| **Overweight** | MR Egger | 14 | 1.11 (0.59-2.08) | 0.32 | 0.75 | 0.07 | 39.3 |
| **Hip circumference** | Inverse variance weighted | 49 | 1.74 (1.34-2.25) | 0.13 | 2.68e-05 | 0.07 | 24.5 |
| **Hip circumference** | Weighted median | 49 | 1.89 (1.33-2.67) | 0.18 | 3.29e-04 | NA | NA |
| **Hip circumference** | MR Egger | 49 | 3.45 (1.35-8.77) | 0.48 | 0.01 | 0.09 | 22.5 |
| **Waist circumference** | Inverse variance weighted | 45 | 1.90 (1.49-2.42) | 0.12 | 1.71e-07 | 0.28 | 10.0 |
| **Waist circumference** | Weighted median | 45 | 1.86 (1.29-2.67) | 0.19 | 8.35e-04 | NA | NA |
| **Waist circumference** | MR Egger | 45 | 2.39 (1.27-4.52) | 0.32 | 0.01 | 0.27 | 10.8 |
| **Waist-to-hip ratio** | Inverse variance weighted | 30 | 1.34 (0.97-1.84) | 0.16 | 0.08 | 0.43 | 2.5 |
| **Waist-to-hip ratio** | Weighted median | 30 | 1.15 (0.72-1.84) | 0.24 | 0.55 | NA | NA |
| **Waist-to-hip ratio** | MR Egger | 30 | 1.61 (0.37-6.95) | 0.75 | 0.53 | 0.38 | 5.7 |
| **Coronary artery disease** | Inverse variance weighted | 61 | 0.99 (0.91-1.08) | 0.04 | 0.79 | 0.65 | 0.00 |
| **Coronary artery disease** | Weighted median | 61 | 0.95 (0.83-1.09) | 0.07 | 0.47 | NA | NA |
| **Coronary artery disease** | MR Egger | 61 | 0.92 (0.77-1.09) | 0.09 | 0.34 | 0.65 | 0.00 |
| **Type 2 diabetes** | Inverse variance weighted | 25 | 1.14 (1.02-1.27) | 0.05 | 0.02 | 0.06 | 32.5 |
| **Type 2 diabetes** | Weighted median | 25 | 1.11 (0.97-1.27) | 0.07 | 0.14 | NA | NA |
| **Type 2 diabetes** | MR Egger | 25 | 1.20 (0.79-1.83) | 0.22 | 0.40 | 0.046 | 35.1 |
| **HDL cholesterol** | Inverse variance weighted | 84 | 0.91 (0.82-1.02) | 0.06 | 0.11 | 0.73 | 0.00 |
| **HDL cholesterol** | Weighted median | 84 | 1.03 (0.87-1.21) | 0.09 | 0.77 | NA | NA |
| **HDL cholesterol** | MR Egger | 84 | 1.16 (0.94-1.43) | 0.11 | 0.17 | 0.87 | 0.00 |
| **Urate** | Inverse variance weighted | 25 | 1.07 (0.94-1.23) | 0.07 | 0.31 | 0.11 | 26.8 |
| **Urate** | Weighted median | 25 | 1.08 (0.90-1.31) | 0.10 | 0.40 | NA | NA |
| **Urate** | MR Egger | 25 | 0.90 (0.71-1.15) | 0.12 | 0.41 | 0.17 | 21.3 |
| **Smoking status: Never** | Inverse variance weighted | 77 | 0.54 (0.27-1.06) | 0.35 | 0.07 | 0.10 | 17.7 |
| **Smoking status: Never** | Weighted median | 77 | 0.84 (0.31-2.24) | 0.50 | 0.72 | NA | NA |
| **Smoking status: Never** | MR Egger | 77 | 1.69 (0.08- 35.31) | 1.55 | 0.74 | 0.09 | 18.2 |
| **Smoking status: Current** | Inverse variance weighted | 15 | 0.94 (0.10-8.84) | 1.15 | 0.96 | 0.95 | 0 |
| **Smoking status: Current** | Weighted median | 15 | 0.24 (0.01-4.54) | 1.50 | 0.34 | NA | NA |
| **Smoking status: Current** | MR Egger | 15 | 4.11 (0.00-1.4e5) | 5.34 | 0.80 | 0.93 | 0 |

**Supplemental Table 12. Egger intercepts for Mendelian Randomization analyses on DKD (all vs. Ctrl phenotype).**

| Exposure | Egger intercept | Intercept SE | Intercept p-value |
|---|---|---|---|
| **Body fat** | 0.00094 | 0.051 | 0.99 |
| **BMI** | -0.0020 | 0.0063 | 0.76 |
| **Obesity class I** | 0.0072 | 0.019 | 0.71 |
| **Obesity class II** | -0.0083 | 0.027 | 0.77 |
| **Overweight** | 0.024 | 0.026 | 0.37 |
| **Waist circumference** | -0.0095 | 0.011 | 0.39 |
| **Hip circumference** | -0.0071 | 0.0093 | 0.45 |
| **WHR** | 0.0057 | 0.017 | 0.74 |
| **CAD** | 0.0086 | 0.0064 | 0.18 |
| **T2D** | 0.012 | 0.012 | 0.31 |
| **HDL cholesterol** | -0.013 | 0.0047 | 0.0085 |
| **Urate** | 0.014 | 0.0090 | 0.12 |
| **Ever smoking** | -0.0084 | 0.013 | 0.51 |
| **Current smoking** | -0.0078 | 0.027 | 0.78 |

**Supplemental Figure 1: Manhattan and QQ-plots of the ten DKD GWAS meta-analysis results.**
**A: Severe DKD** (Macroalbuminuria or ESRD vs. normal AER). $\lambda_{GC}$ = 1.029, LD score regression (LDSR) intercept = 1.019.

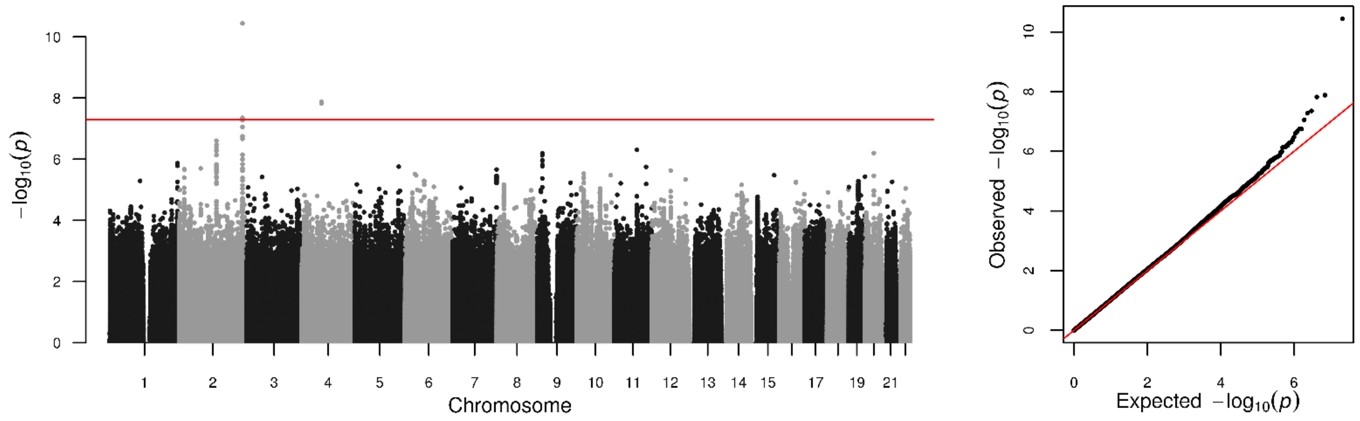

**B: Macro** (Macroalbuminuria vs. normal AER). $\lambda_{GC}$ = 1.002, LDSR intercept = 1.028.

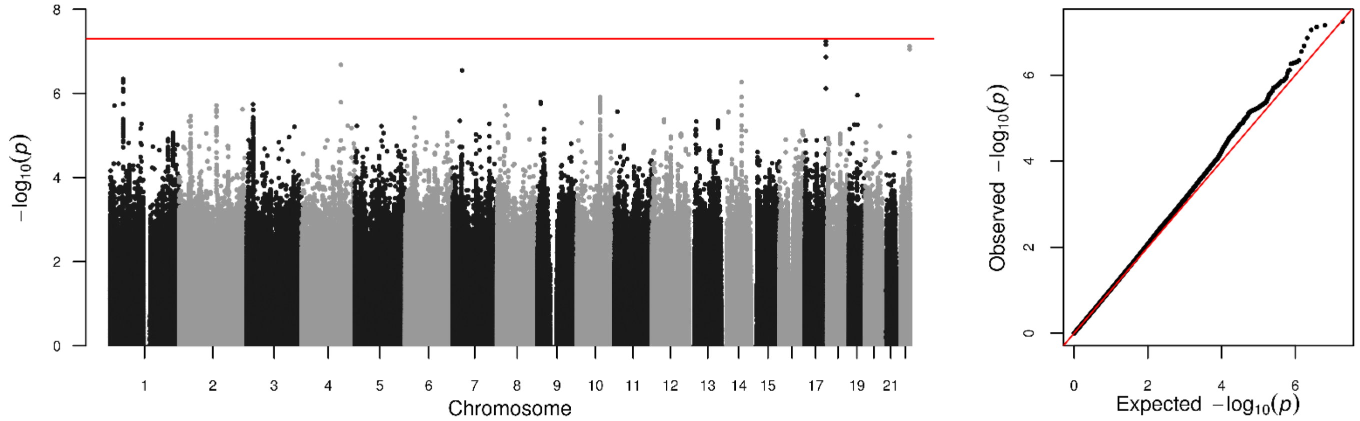

**C: ESRD** (ESRD vs. normal AER). $\lambda_{GC}$ = 1.011, LDSR intercept = 1.018.

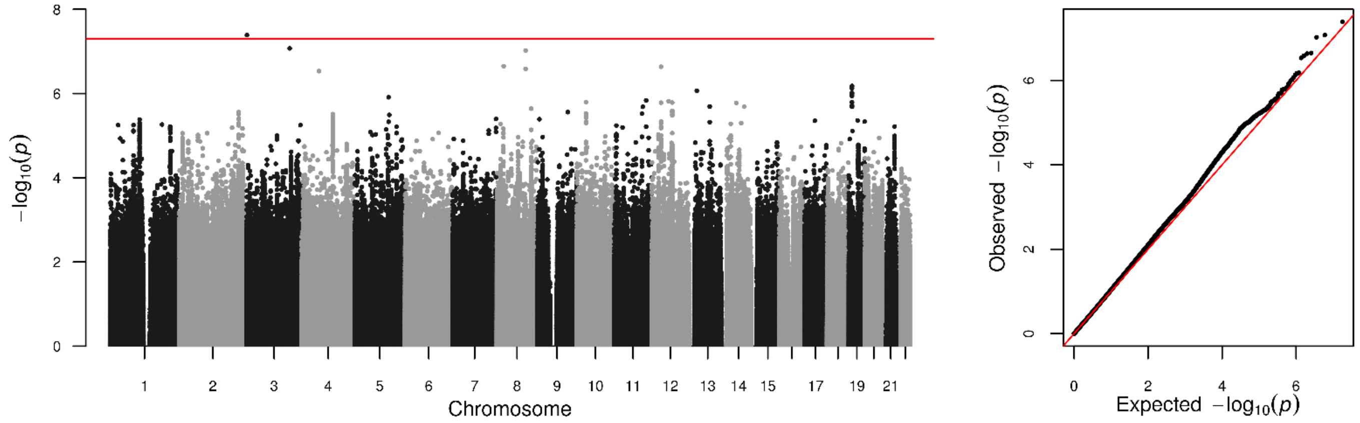

**D: ESRD vs. All** (ESRD vs. macro- or microalbuminuria or normal AER). $\lambda_{GC}$ = 1.029, LDSR intercept = 1.054.

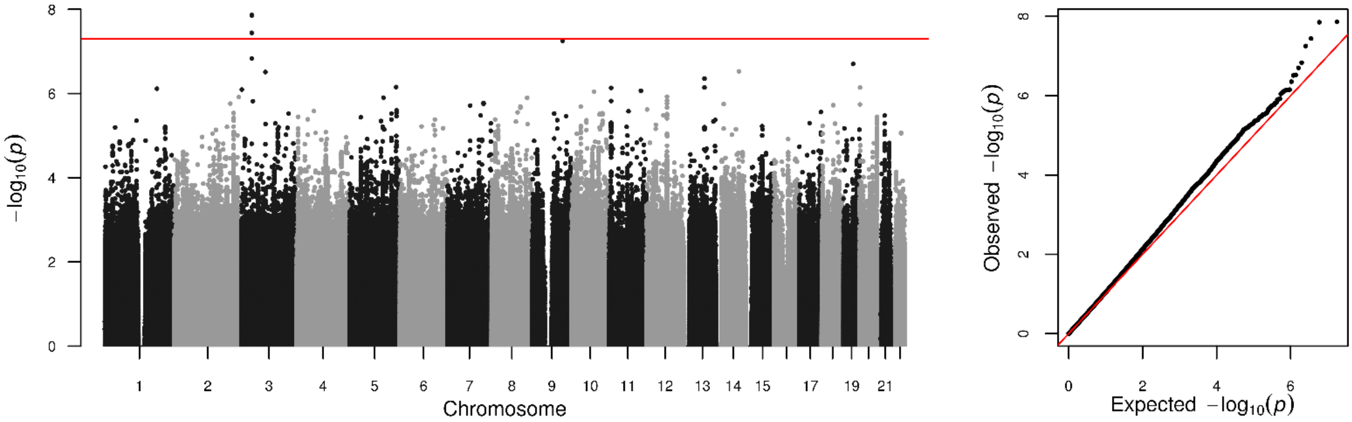

**E: ESRD vs. Macro** (ESRD vs. macroalbuminuria). $\lambda_{GC}$ = 1.011, LDSR intercept = 1.009.

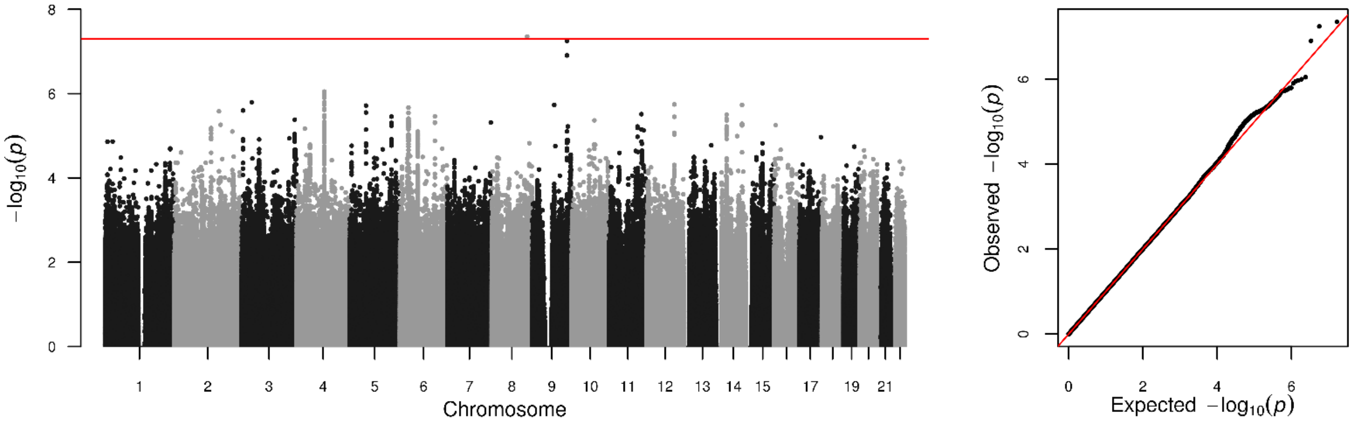

**F: All vs. Ctrl** (Micro- or Macroalbuminuria or ESRD vs. normal AER). $\lambda_{GC}$ = 1.035, LDSR intercept = 1.005.

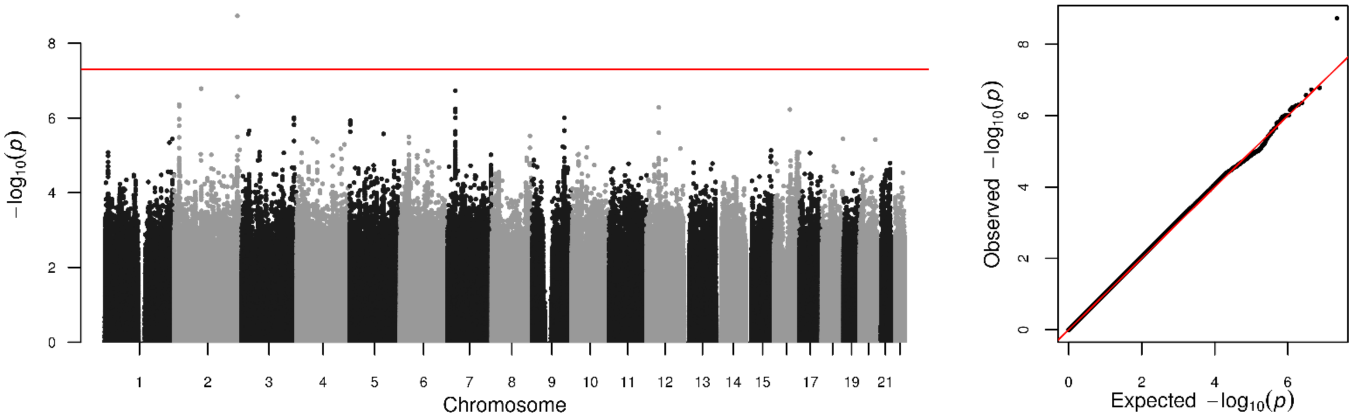

**G: Micro** (Microalbuminuria vs. normal AER). $\lambda_{GC}$ = 1.008, LDSR intercept = 1.004.

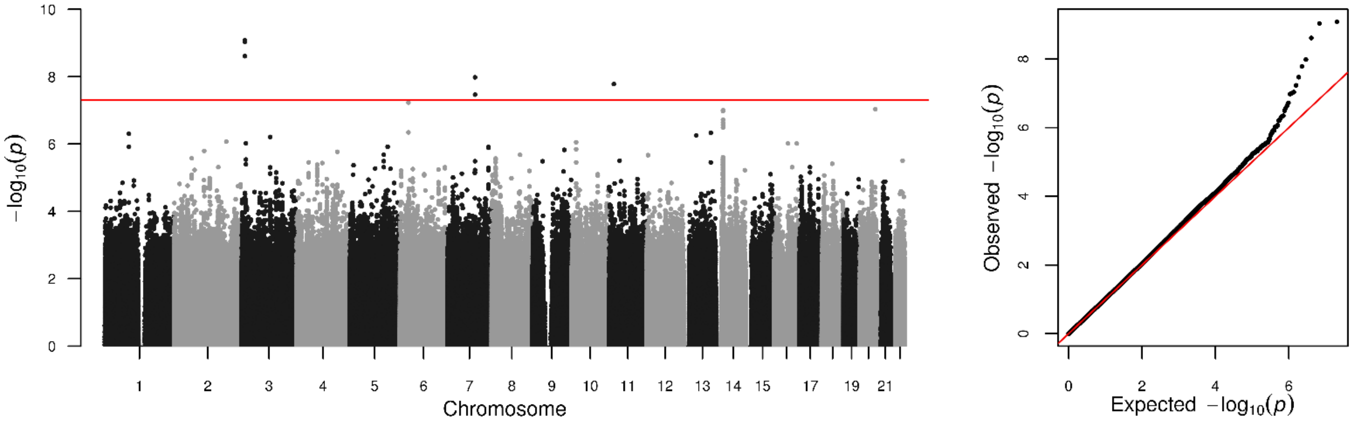

**H: CKD** (eGFR < 60 ml/min/1.73m$^2$ vs. eGFR ≥60 ml/min/1.73m$^2$). $\lambda_{GC}$ = 1.041, LDSR intercept = 1.028.

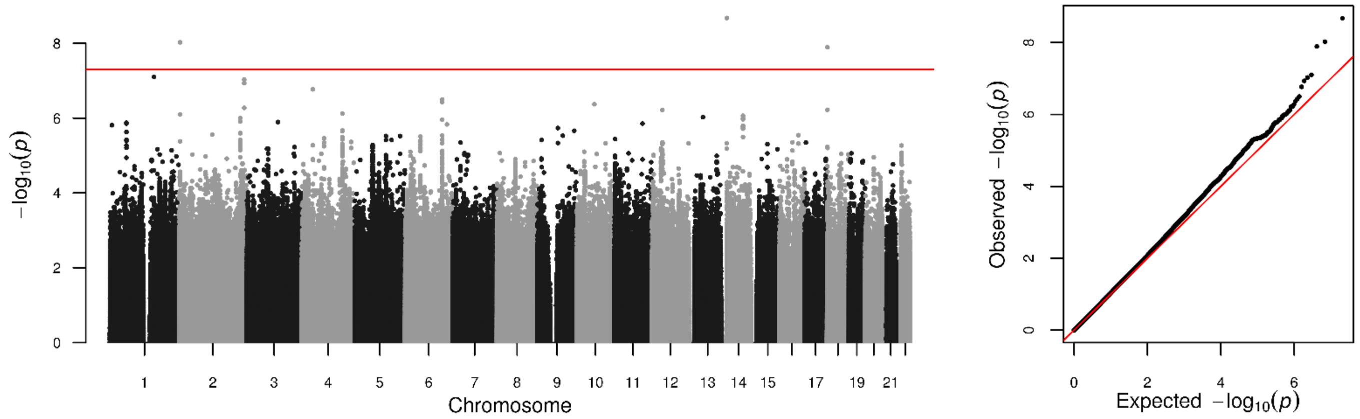

**I: CKD Extremes** (ESRD or eGFR < 15 ml/min/1.73m$^2$ vs. eGFR ≥ 60 ml/min/1.73m$^2$). $\lambda_{GC}$ = 1.023, LDSR intercept = 1.019.

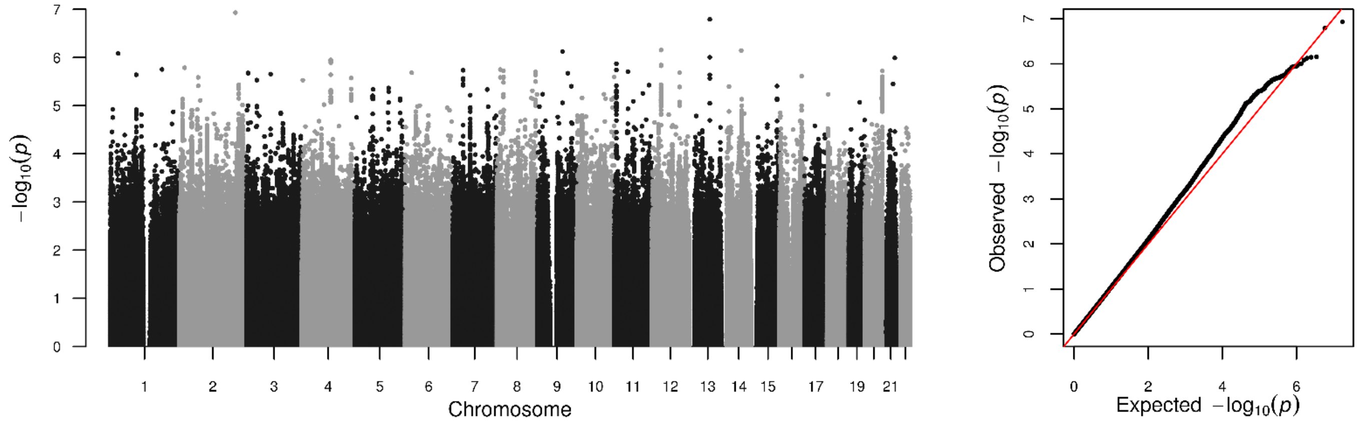

**J: CKD-DKD** (ESRD or eGFR < 45 ml/min/1.73m$^2$ and micro- or macroalbuminuria vs. eGFR ≥ 60ml/min/1.73m$^2$ and normal AER). $\lambda_{GC}$ = 1.023, LDSR intercept = 1.031.

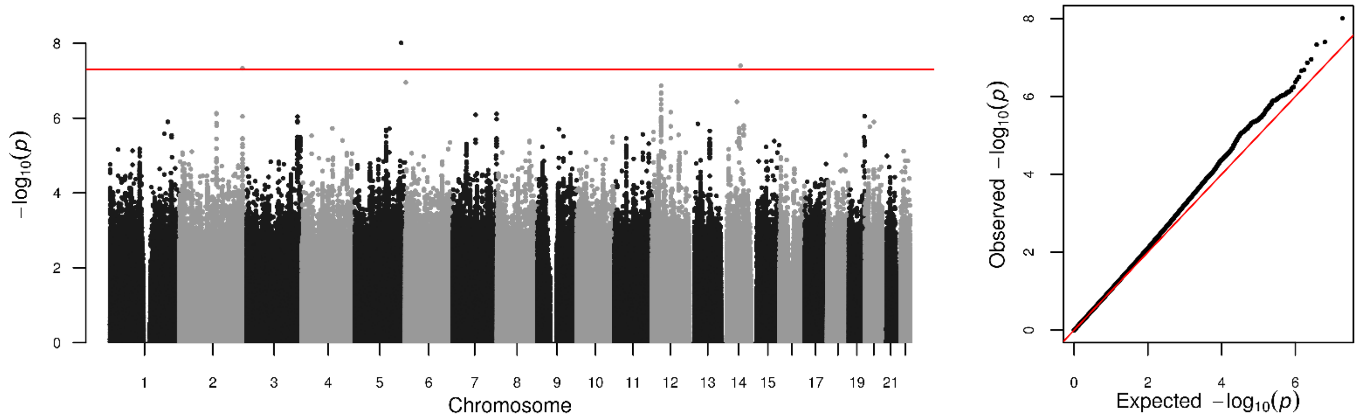

**Supplemental Figure 2: Regional association plots for the GWAS meta-analysis lead loci (A-K).**

**A:** CKD+DKD chr5:166978230 (rs72831309)

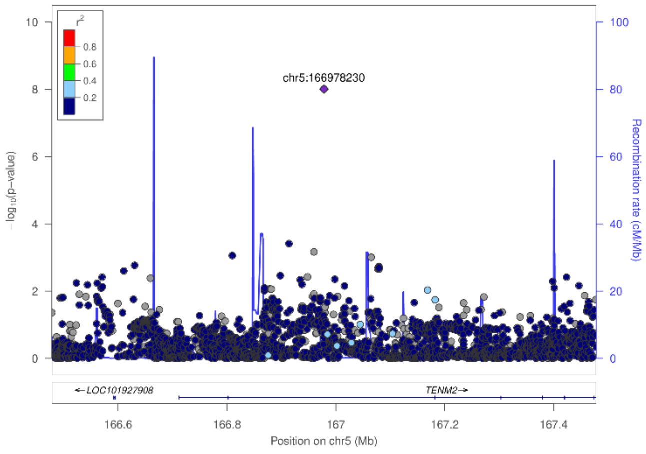

**B:** CKD chr2:3745215 (rs12615970)

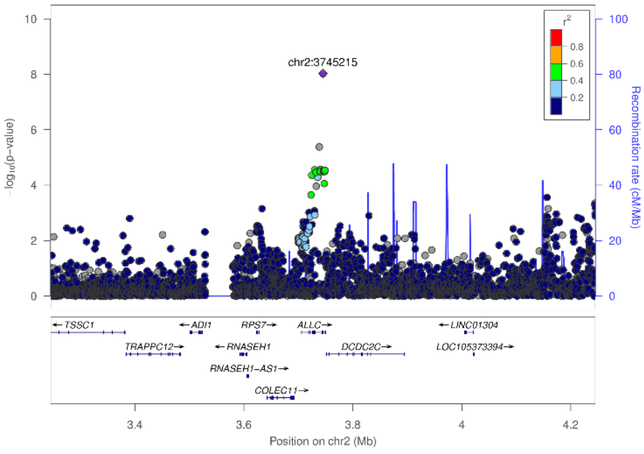

**C:** Severe DKD chr2:228121101 (rs55703767)

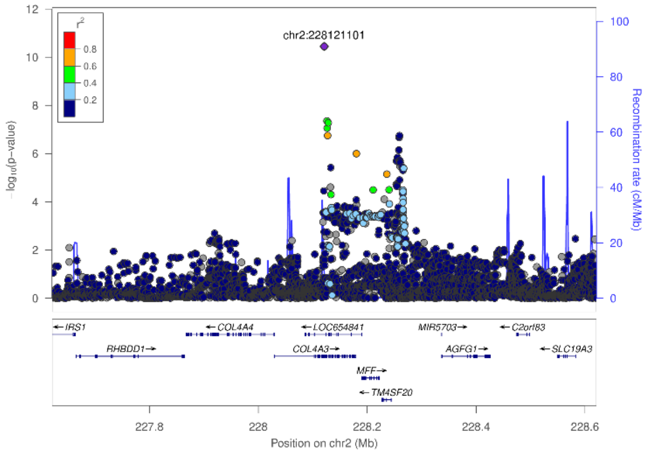

**D:** ESRD chr3:926345 (rs115061173)

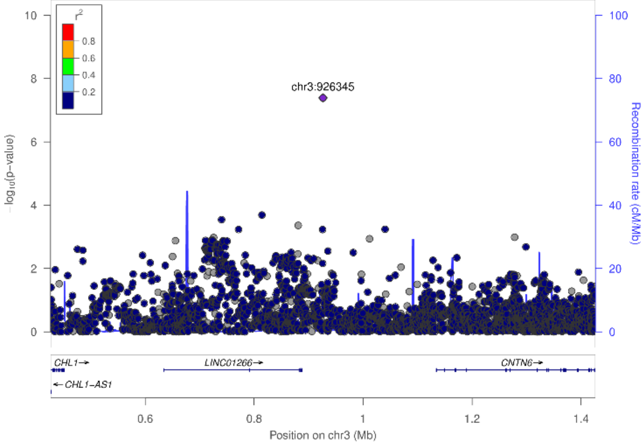

**E:** Micro chr3:11910635 (rs142823282)

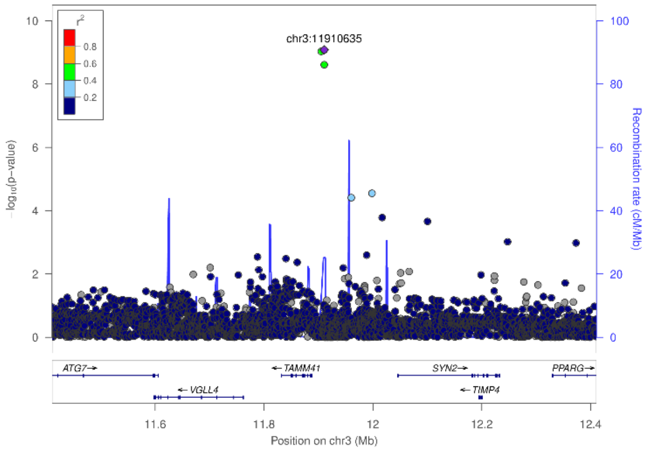

**F:** ESRD vs. All chr3:36566312 (rs116216059). The SNP rs116216059 is located on a single nucleus ATACseq (snATACseq) peak border in podocytes (PODO), peak value 1.1 (peak maximum value 7.0).

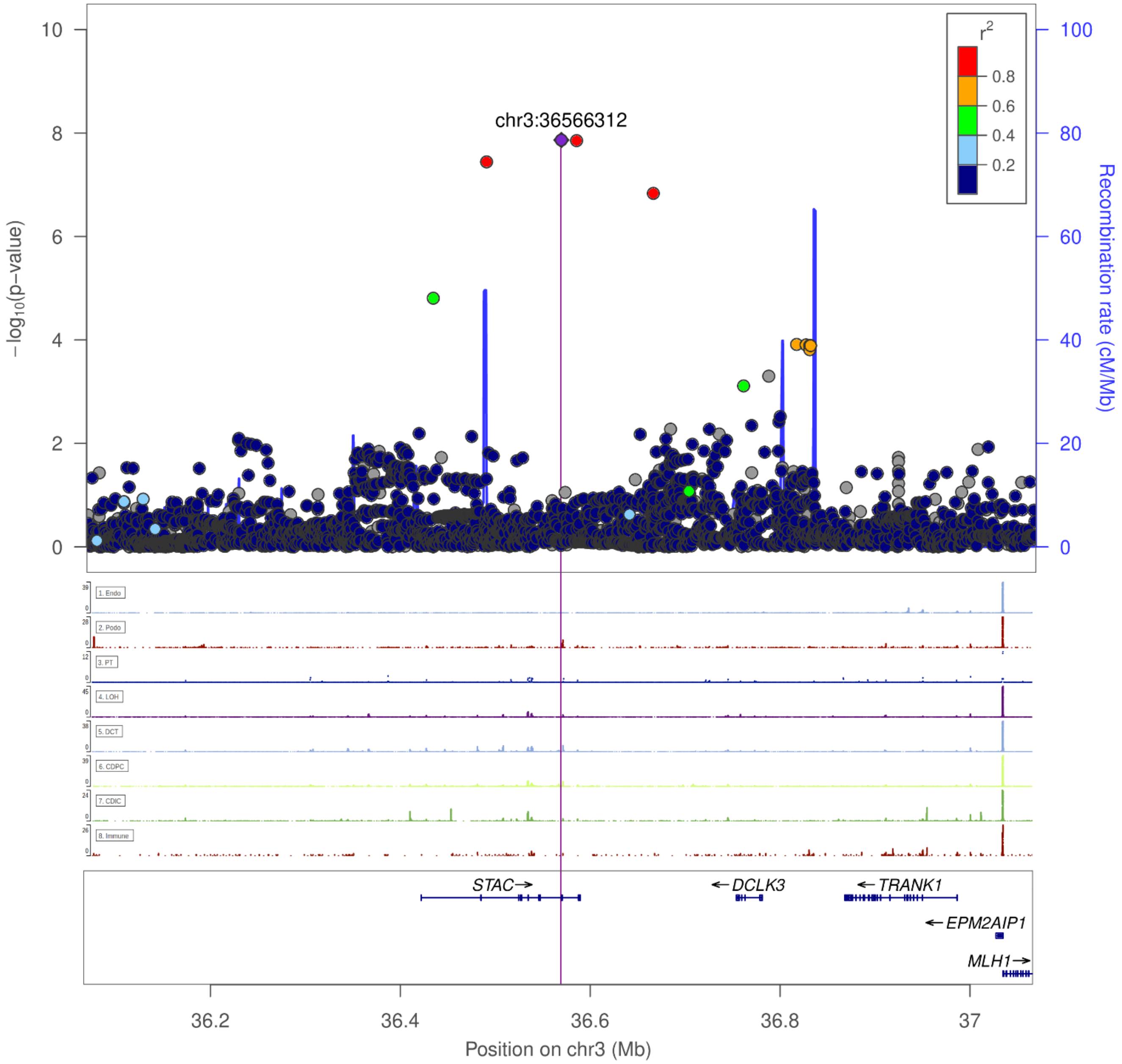

**G:** Severe DKD chr4:71358776 (rs191449639)

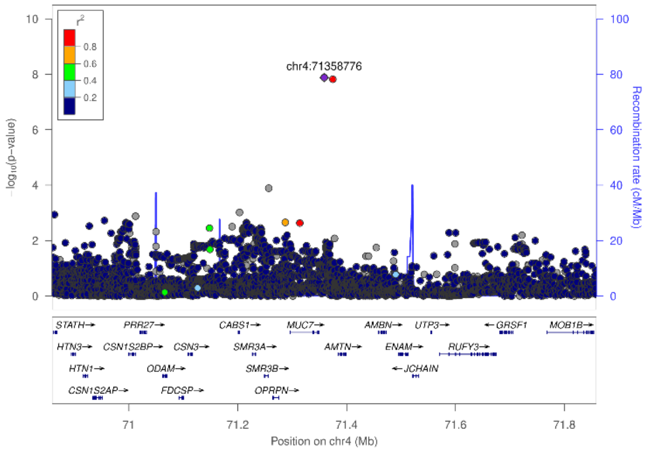

**H:** Micro chr7:99728546 (rs77273076)

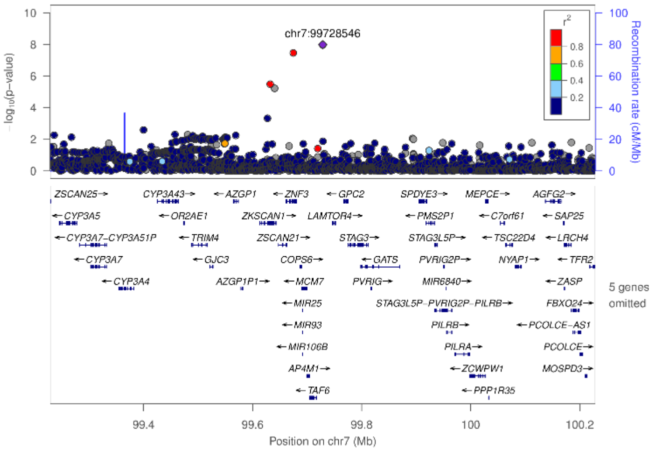

**I:** ESRD vs. macro chr8:128100029 (rs551191707)

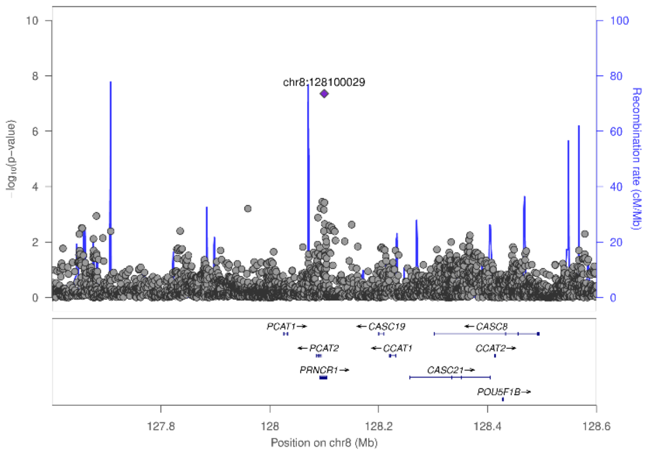

**J:** Micro chr11:16937846 (rs183937294)

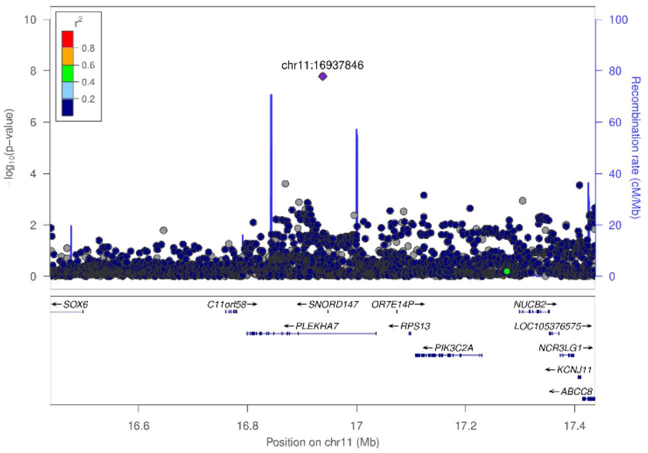

**K:** CKD chr18:1811108 (rs185299109)

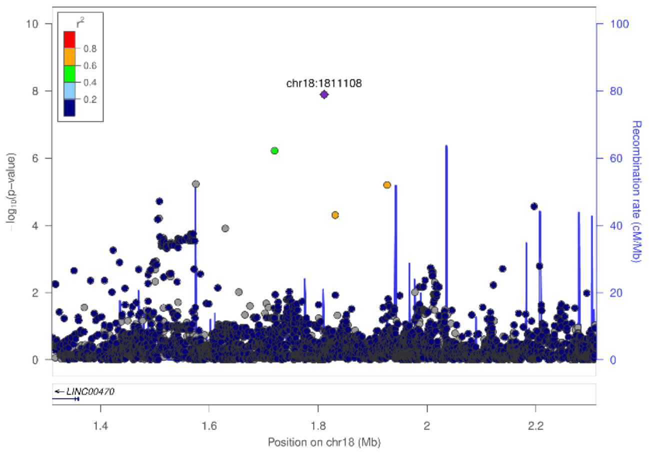

**Supplemental Figure 3: Regional association plot for the *COL4A3* gene region associated with Severe DKD, indicating a secondary association peak at chr2:228259302 (rs6436688, effect allele (A) frequency 56%, OR = 1.13 (95% confidence interval 1.08 – 1.19), p-value 1.79x10$^{-7}$).** SNP rs6436688 is in partial LD (D'=0.51, $r^2$=0.08, 1000Genomes European ancestry populations) with the original *COL4A3* lead variant rs55703767. Variants are colored according to their LD correlation with the primary signal (chr2:228121101 rs55703767) in red, or with the secondary peak in blue; stronger color indicates stronger correlation.

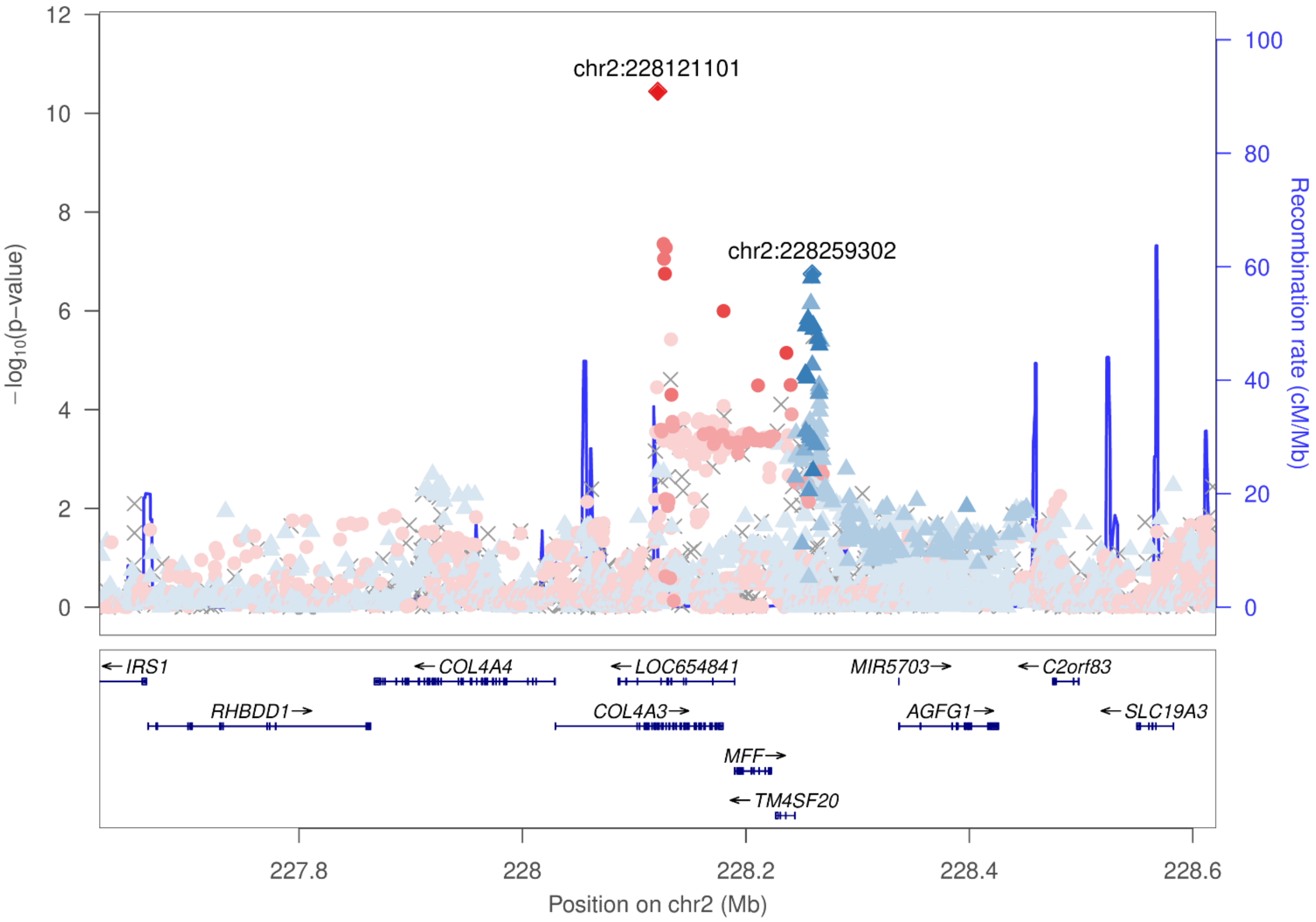

**Supplemental Figure 4: Regional association plots for the gene-level analysis results from MAGMA and PASCAL analysis.** The implicated gene region is highlighted in light blue. If the same gene was significant in both analyses, only MAGMA region is highlighted (gene flanking ± 50 kbp, vs. ± 5 kbp for PASCAL).

**A:** CKD, *PTPRN*

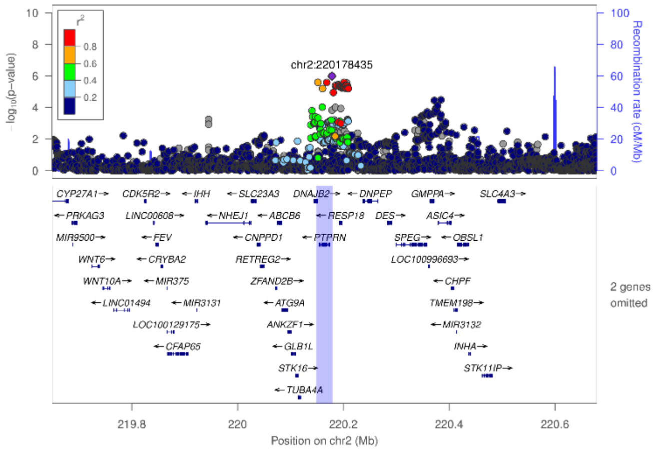

**B:** CKD, *RESP18*

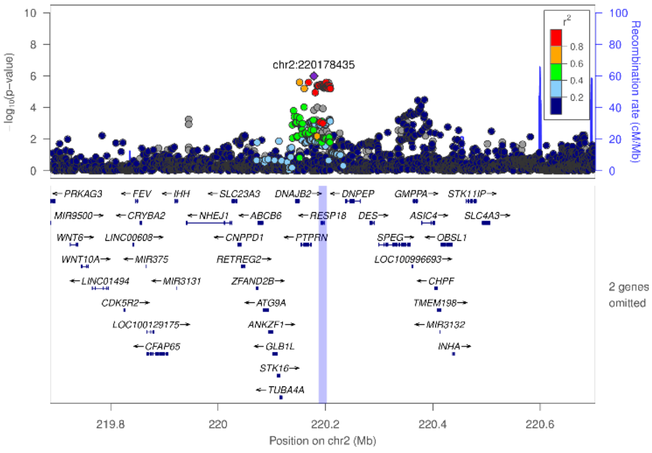

**C:** Severe DKD, *MFF*

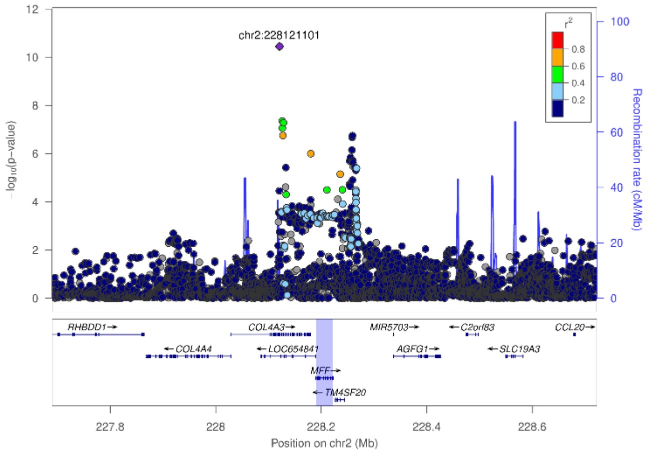

**D:** ESRD vs. macro, *EIF4E*

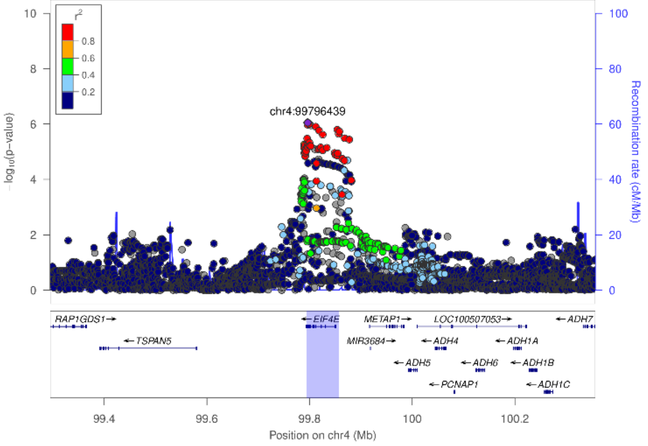

**E:** DKD, *INIP*

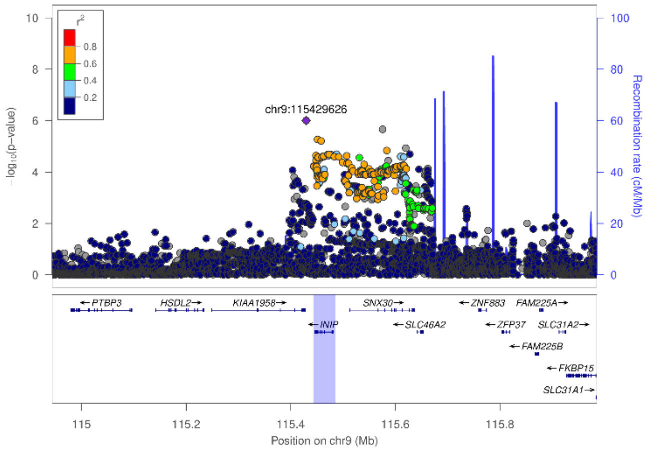

**F:** DKD, *SNX30*

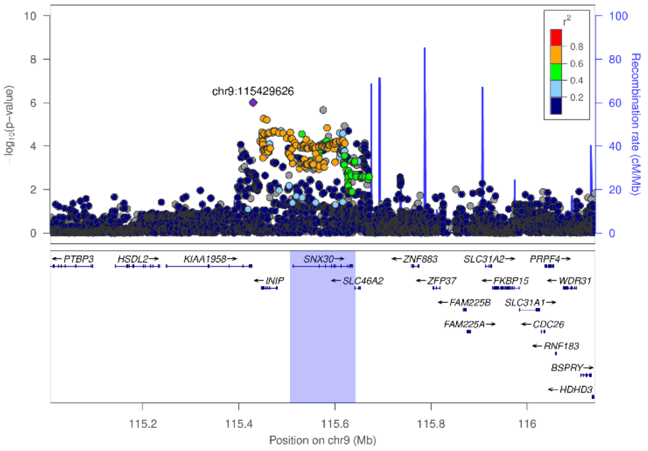

**G:** Severe DKD, *GPR158*

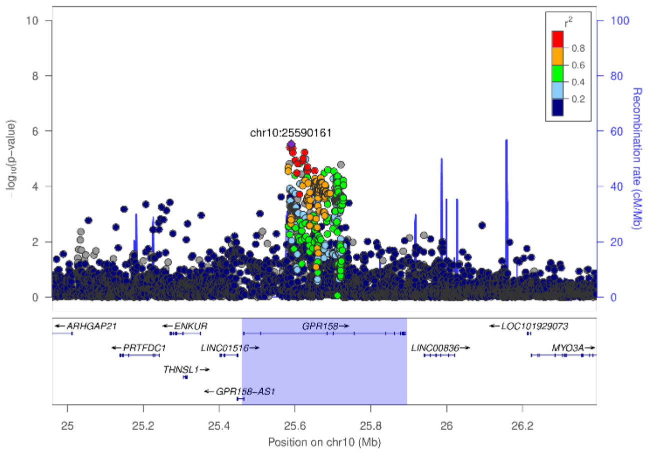

**H:** ESRD vs. macro, DCLK1

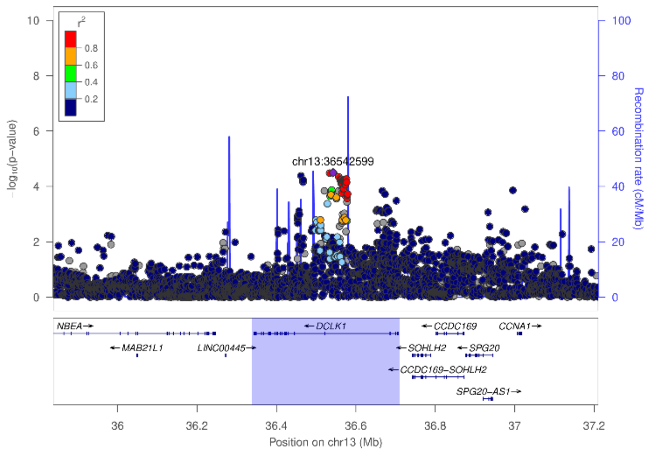

**I:** Severe DKD, *LSM14A*

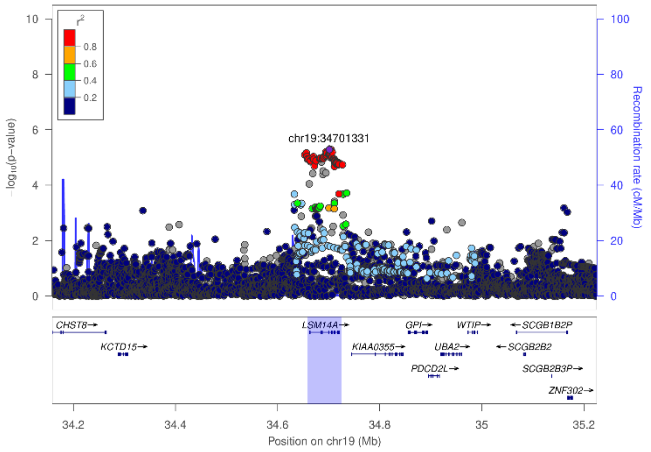

**J:** CKD extremes, COL20A1

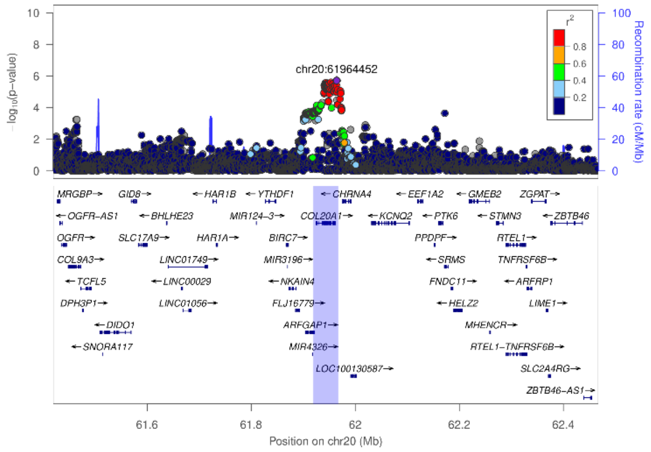

**Supplemental Figure 5: Gene prioritization for the COL4A3 gene at lead SNP rs5570367 associated with Severe DKD using multiple intersecting gene prioritization approaches (PoPS, nearest gene, and MAGMA).** Plotted is the PoP Score (y-axis) versus the genes within a 500kb flanking window surrounding the lead SNP (x-axis), colored by distance to the lead SNP, and bolded if the gene was also within the top 10% of prioritized genes genome-wide using MAGMA.

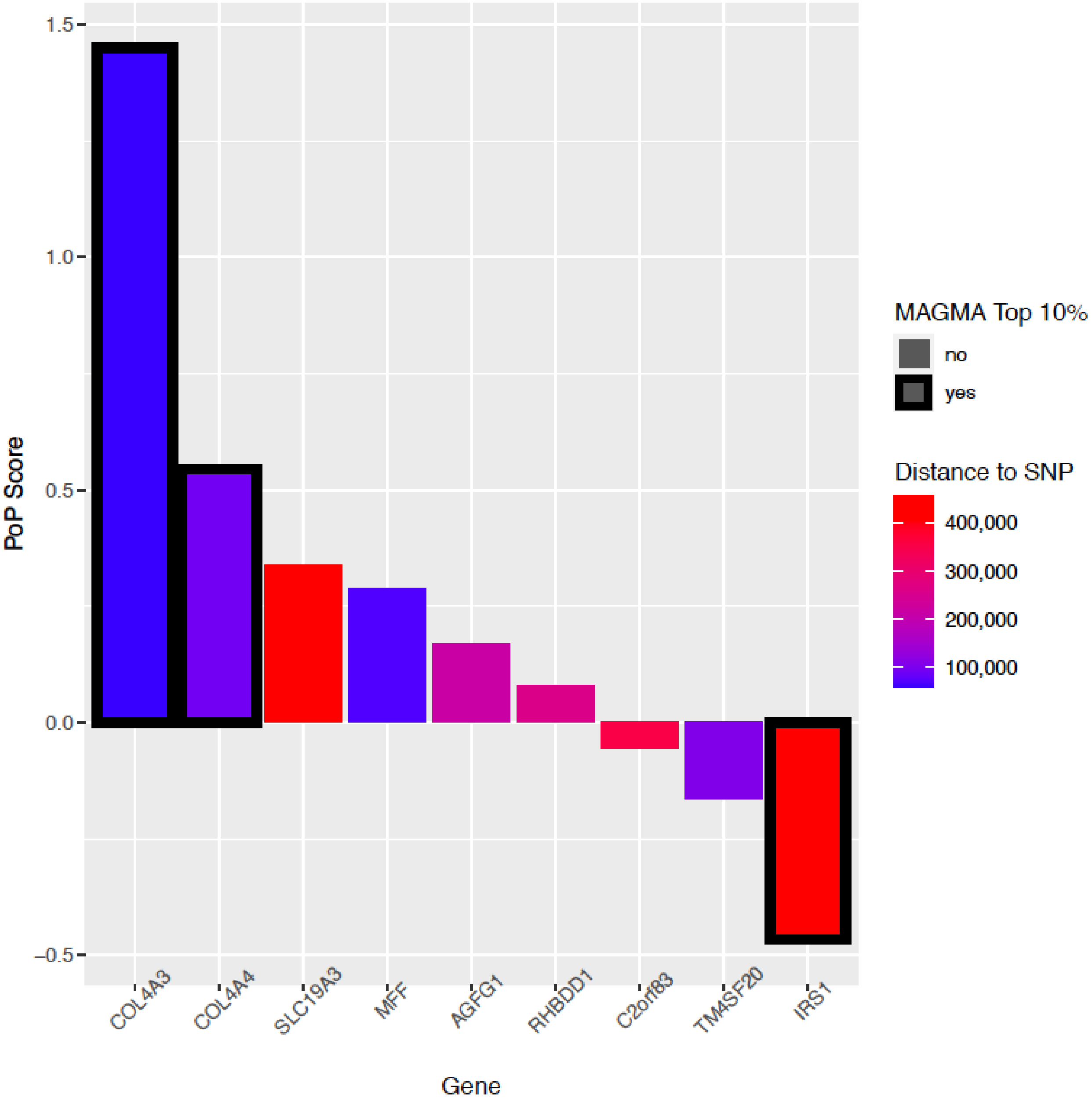

**Supplemental Figure 6: Tubular and glomerular gene expression of the lead genes correlate with multiple morphological and pathological renal parameters.** Golden rectangles indicate glomerular gene expression, green ellipses tubular gene expression, and gray circles the morphological phenotypes. All nominally significant correlations are shown. Blue edges indicate negative correlation, red edges positive correlation. Correlation with fibrosis, Glomerulosclerosis (GlomScl), and eGFR are measured in the nephrectomy samples. B1_ and B2_ indicate phenotypes from the first and second renal biopsies (B1, B2, respectively) from the Pima Indians, correlated with gene expression in transcriptomic data from the corresponding time point. B1/2_GlomVol: glomerular volume; B1/2_GlomW: glomerular width; B1/2 FPW: podocyte foot process width. B1/2_ACR: albumin creatinine ratio; B1/2_Fibr: fibrosis; B1/2_HbA1c: HbA1c B1/2_MesVol: mesangial volume; B1/2_SV: Surface volume of peripheral glomerular basement membrane per glomerulus; B1_Slope: measured GFR (mGFR) slope between the B1 and B2. B1_DMesVol: change in mesangial volume between B1 and B2. B1_DSV: change in SV between B1 and B2.

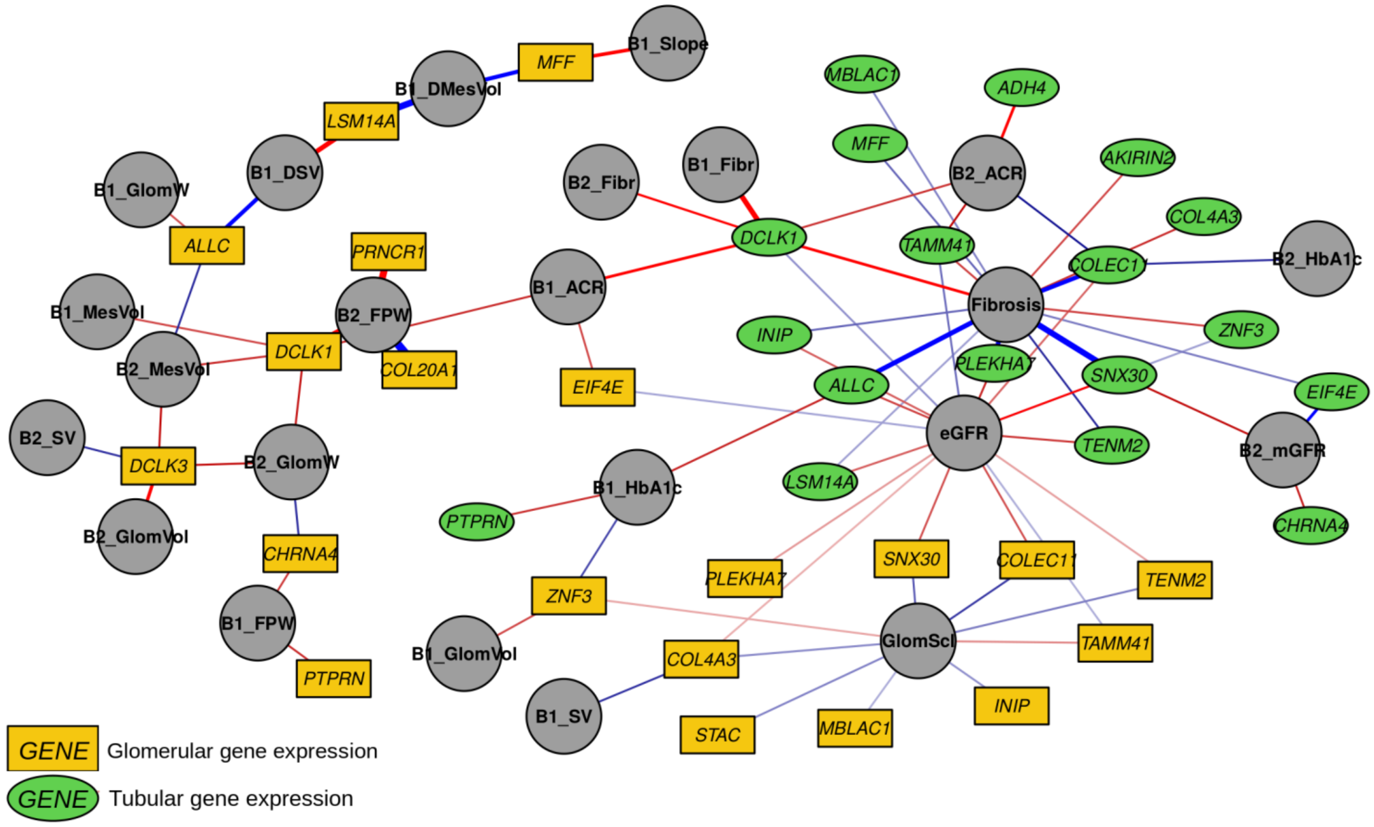

**Supplemental Figure 7: Genetic correlation between DKD and related traits based on LD score regression.** Only trait combinations with p<0.05 are shown, and traits that remained significant after correcting for 78 studied traits (p-value < $0.05/78 = 6.4\times10^{-4}$) are indicated with dark dot borders. Dot colors indicate aging related (light purple), anthropometric (green; including BMI and obesity related (light green), height (dark green), and waist and/or hip related (pale green)), inflammatory bowel disease (orange), bone mineral density (light gray), coronary artery disease (red), glycemic (light blue), type 2 diabetes (dark blue), serum creatinine and cystatin C (brown), lipids (yellow), uric acid (purple), and smoking related (dark gray) traits.

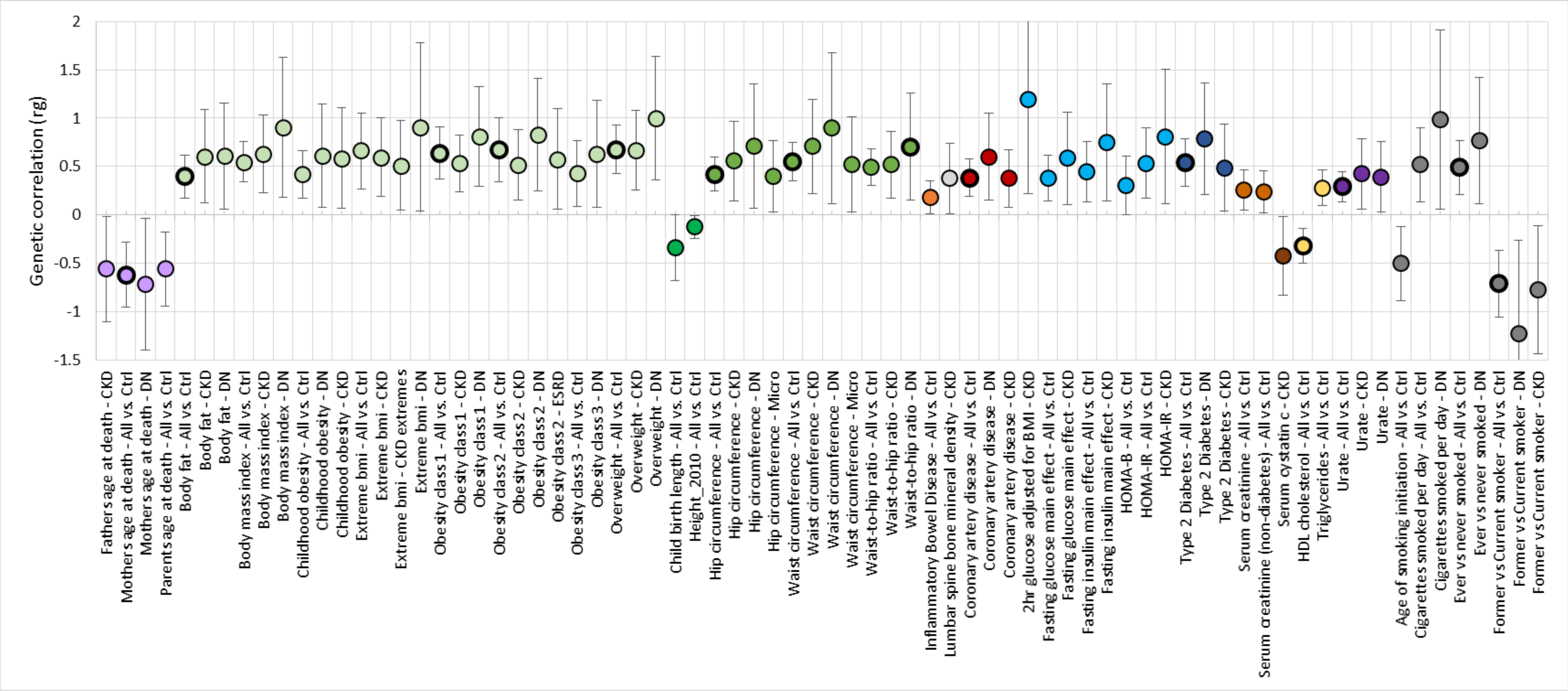

**Supplemental Figure 8: Mendelian Randomization scatter plots for SNP effects for the metabolic traits vs. DKD (All vs. Ctrl).** Lines indicate IVW, Weighted median, and MR egger coefficients.

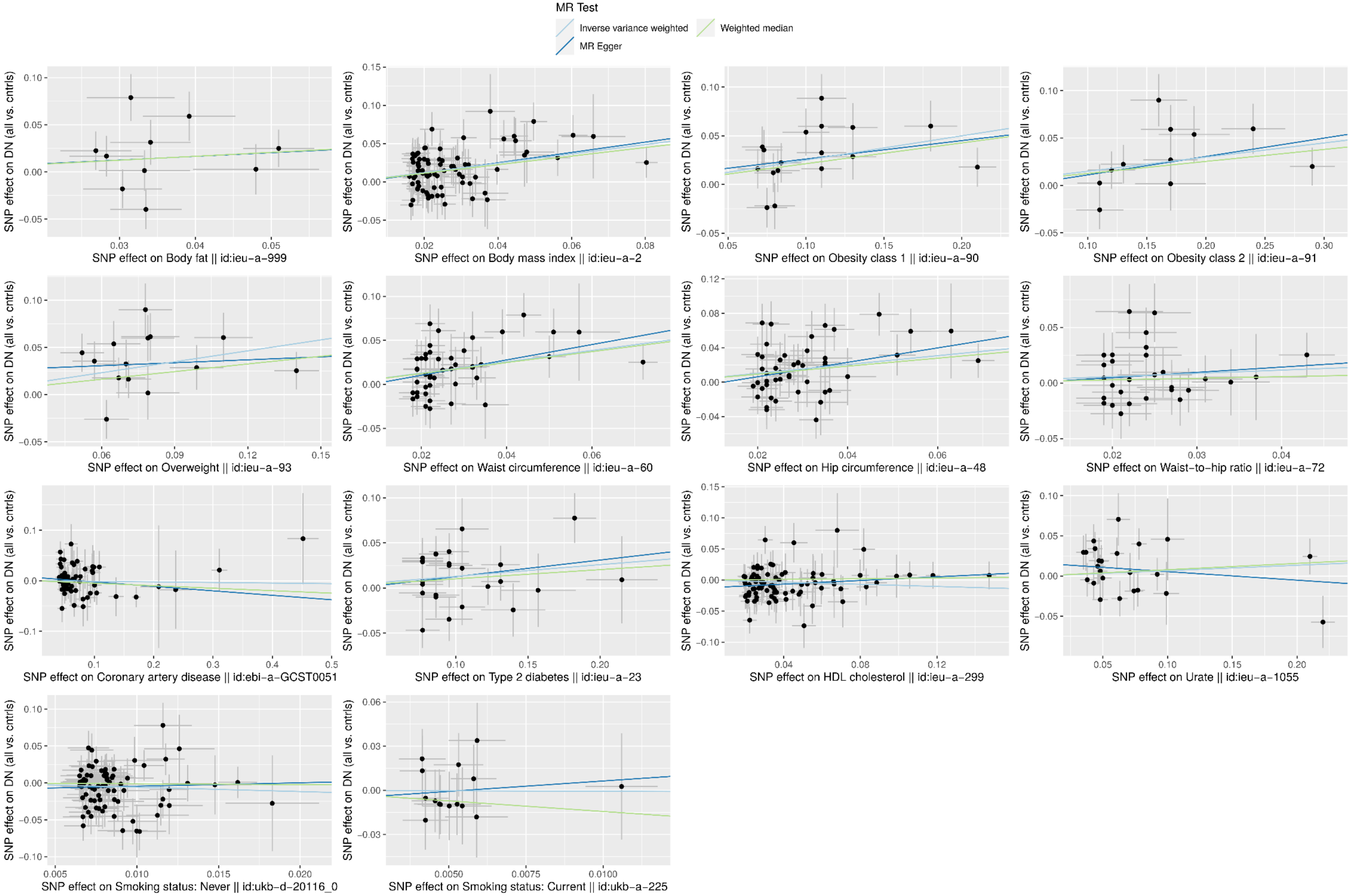

**Supplemental Figure 9: rs1260634 intronic in the ALLC gene affects the predicted binding motifs for KLF12, KLF4, and SP8 (top to bottom).** Images obtained from the RegulomeDB (www.regulomedb.org)

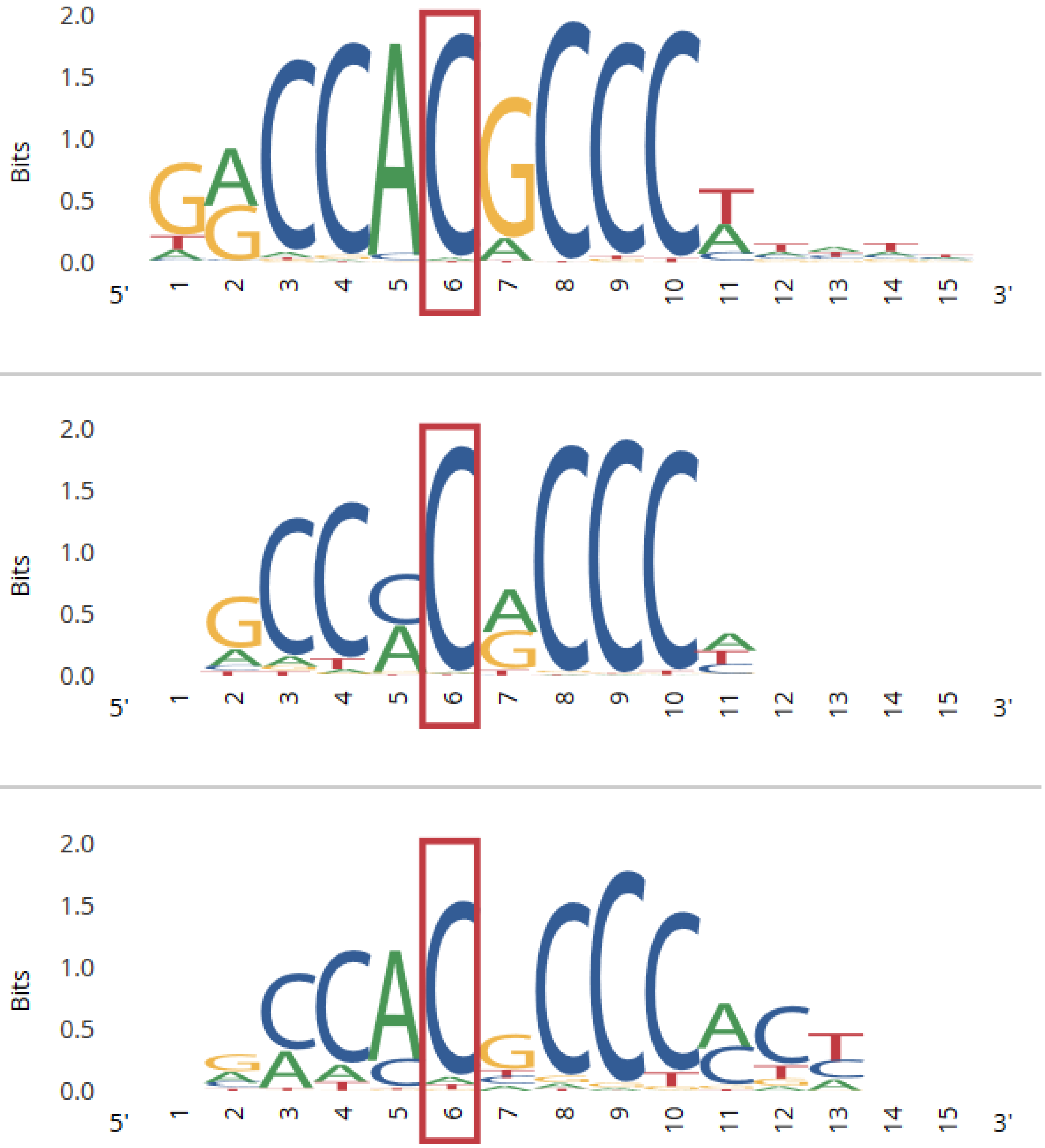

STROBE Statement—checklist of items that should be included in reports of observational studies

| | Item No | Recommendation | Page No |
|---|---|---|---|
| **Title and abstract** | 1 | (*a*) Indicate the study's design with a commonly used term in the title or the abstract | 1 |
| | | (*b*) Provide in the abstract an informative and balanced summary of what was done and what was found | 1 |
| **Introduction** | | | |
| Background/rationale | 2 | Explain the scientific background and rationale for the investigation being reported | 3 |
| Objectives | 3 | State specific objectives, including any prespecified hypotheses | 3 |
| **Methods** | | | |
| Study design | 4 | Present key elements of study design early in the paper | Fig 1 |
| Setting | 5 | Describe the setting, locations, and relevant dates, including periods of recruitment, exposure, follow-up, and data collection | 4 |
| Participants | 6 | (*a*) *Cohort study*—Give the eligibility criteria, and the sources and methods of selection of participants. Describe methods of follow-up<br>*Case-control study*—Give the eligibility criteria, and the sources and methods of case ascertainment and control selection. Give the rationale for the choice of cases and controls<br>*Cross-sectional study*—Give the eligibility criteria, and the sources and methods of selection of participants | 4 |
| | | (*b*) *Cohort study*—For matched studies, give matching criteria and number of exposed and unexposed<br>*Case-control study*—For matched studies, give matching criteria and the number of controls per case | |
| Variables | 7 | Clearly define all outcomes, exposures, predictors, potential confounders, and effect modifiers. Give diagnostic criteria, if applicable | 4 |
| Data sources/ measurement | 8* | For each variable of interest, give sources of data and details of methods of assessment (measurement). Describe comparability of assessment methods if there is more than one group | Table S1 |
| Bias | 9 | Describe any efforts to address potential sources of bias | 10 |
| Study size | 10 | Explain how the study size was arrived at | Table S2 |
| Quantitative variables | 11 | Explain how quantitative variables were handled in the analyses. If applicable, describe which groupings were chosen and why | Table S1 |
| Statistical methods | 12 | (*a*) Describe all statistical methods, including those used to control for confounding | 5-10 |
| | | (*b*) Describe any methods used to examine subgroups and interactions | |
| | | (*c*) Explain how missing data were addressed | |
| | | (*d*) *Cohort study*—If applicable, explain how loss to follow-up was addressed<br>*Case-control study*—If applicable, explain how matching of cases and controls was addressed<br>*Cross-sectional study*—If applicable, describe analytical methods taking account of sampling strategy | |
| | | (*e*) Describe any sensitivity analyses | |

| **Results** | | | |
|---|---|---|---|
| Participants | 13* | (a) Report numbers of individuals at each stage of study—eg numbers potentially eligible, examined for eligibility, confirmed eligible, included in the study, completing follow-up, and analysed | Table S2 |
| | | (b) Give reasons for non-participation at each stage | |
| | | (c) Consider use of a flow diagram | |
| Descriptive data | 14* | (a) Give characteristics of study participants (eg demographic, clinical, social) and information on exposures and potential confounders | ref to orig papers |
| | | (b) Indicate number of participants with missing data for each variable of interest | |
| | | (c) *Cohort study*—Summarise follow-up time (eg, average and total amount) | |

| | | | |
|---|---|---|---|
| Outcome data | 15* | *Cohort study*—Report numbers of outcome events or summary measures over time | |
| | | *Case-control study*—Report numbers in each exposure category, or summary measures of exposure | Table S2 |
| | | *Cross-sectional study*—Report numbers of outcome events or summary measures | |
| Main results | 16 | (*a*) Give unadjusted estimates and, if applicable, confounder-adjusted estimates and their precision (eg, 95% confidence interval). Make clear which confounders were adjusted for and why they were included | Table 1; Bonf thresholds are given |
| | | (*b*) Report category boundaries when continuous variables were categorized | |
| | | (*c*) If relevant, consider translating estimates of relative risk into absolute risk for a meaningful time period | |
| Other analyses | 17 | Report other analyses done—eg analyses of subgroups and interactions, and sensitivity analyses | |
| **Discussion** | | | |
| Key results | 18 | Summarise key results with reference to study objectives | 15 |
| Limitations | 19 | Discuss limitations of the study, taking into account sources of potential bias or imprecision. Discuss both direction and magnitude of any potential bias | 19 |
| Interpretation | 20 | Give a cautious overall interpretation of results considering objectives, limitations, multiplicity of analyses, results from similar studies, and other relevant evidence | 15-16 |
| Generalisability | 21 | Discuss the generalisability (external validity) of the study results | 19 |
| **Other information** | | | |
| Funding | 22 | Give the source of funding and the role of the funders for the present study and, if applicable, for the original study on which the present article is based | 20 |

*Give information separately for cases and controls in case-control studies and, if applicable, for exposed and unexposed groups in cohort and cross-sectional studies.

**Note:** An Explanation and Elaboration article discusses each checklist item and gives methodological background and published examples of transparent reporting. The STROBE checklist is best used in conjunction with this article (freely available on the Web sites of PLoS Medicine at http://www.plosmedicine.org/, Annals of Internal Medicine at http://www.annals.org/, and Epidemiology at http://www.epidem.com/). Information on the STROBE Initiative is available at www.strobe-statement.org.
